# Heterogeneous Treatment Effects in HFpEF: Distinguishing Drug-Specific Response from Prognostic Phenotypes Across Randomized Trials

**DOI:** 10.64898/2026.07.06.26357251

**Authors:** Clodomir Santana, Asuka Katayama, Aditya Ballal, Padmini Sirish, David A. Liem, Julie T. Bidwell, Chao-Yin Chen, Miriam Nuño, Imo A. Ebong, Xiao-Dong Zhang, Leighton Izu, Laura Alagna, Lorenzo L. Pesce, Ryan K. Sisk, Barry A. Borlaug, Javed Butler, Julio A. Chirinos, William A. Chutkow, Akshay S. Desai, Patrice Desvigne-Nickens, Michael M. Givertz, Sadiya S. Khan, Dalane W. Kitzman, Gregory D. Lewis, Laura J. Rasmussen-Torvik, Margaret M. Redfield, Svati H. Shah, Kavita Sharma, Jennifer L. Taylor, Emily Tinsley, Renee Wong, Sanjiv J. Shah, Javier E. López, Nipavan Chiamvimonvat, Martin Cadeiras

## Abstract

**Background:** Heart failure with preserved ejection fraction (HFpEF) is a heterogeneous syndrome comprising multiple pathophysiological phenotypes. HFpEF trials have largely enrolled diverse populations and reported average treatment effects, consistently yielding neutral results that may obscure drug-specific benefits within distinct subgroups. To address this issue, we employ an interaction-based that incorporates treatment-by-variable interactions to uncover drug-specific responses.

**Methods:** We leveraged four HFpEF clinical trials (TOPCAT, RELAX, NEAT-HFpEF, INDIE-HFpEF) and developed a framework comprising two complementary approaches. The first employed a prognostic responder model to evaluate whether conventional responder definitions reflect treatment-specific benefit or instead capture favorable clinical trajectories common to both treatment and placebo groups. The second used an interaction-based individual treatment effect (ITE) modeling to identify baseline variables that modify therapy effect, distinguishing drug-specific response from prognostic phenotypes.

**Results:** Although the prognostic responder model demonstrated good discrimination, further analisys suggested it primarily captured a prognostic signal associated with favorable clinical trajectories common to both treatment and placebo arms. In contrast, the ITE model identified distinct, drug-specific effect modifiers across trials (cardiorenal-inflammatory for spironolactone (TOPCAT), NO-mediated anti-inflammatory for isosorbide mononitrate (NEAT-HFpEF), afterload-reducing for inorganic nitrite (INDIE-HFpEF), and anti–volume-overload for sildenafil (RELAX). Each ITE model demonstrated significance only within its own trial suggesting drug-specific signal.

**Conclusions:** The proposed method identifies mechanism-specific effect modifiers, and uncovers clinically meaningful heterogeneity in treatment response, which is not captured by conventional MCID-based approaches. Although exploratory, these findings support phenotype-guided therapy in HFpEF and argue for phenotype-informed trial design to enhance treatment-effect detection and therapy targeting.

**Graphical Abstract.:** 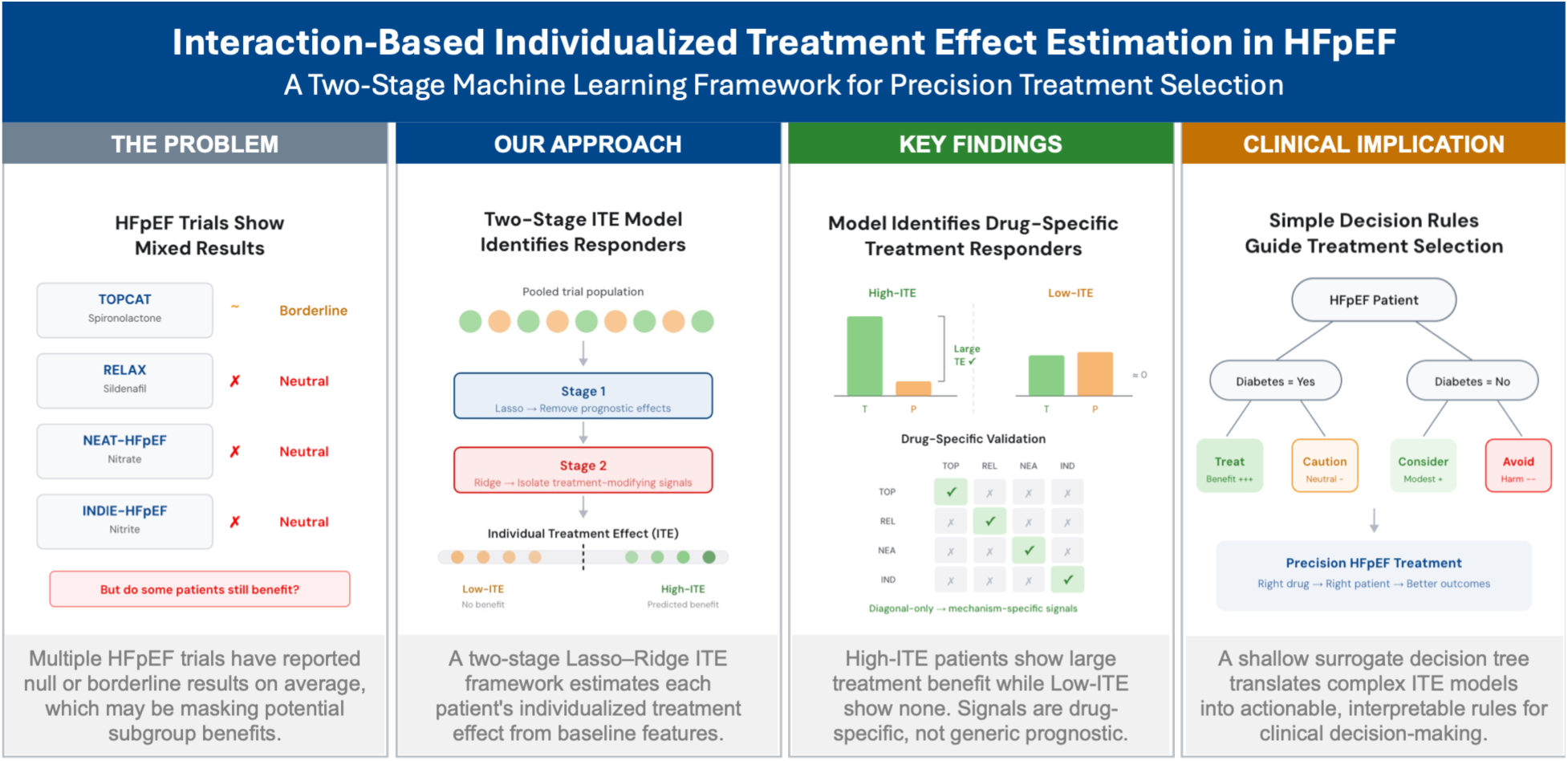

## 1. Introduction

Heart failure with preserved ejection fraction (HFpEF) accounts for approximately half of all heart failure cases^1^, and represents a major and growing public health problem burden, driven by increasing prevalence, substantial morbidity and mortality, and limited treatment options^2,3^. Indeed, HFpEF has been recognized as one of the most urgent unmet needs in cardiovascular medicine^4^. HFpEF is a systemic, multiorgan, and progressive syndrome characterized by marked phenotypic heterogeneity and a high burden of comorbidities, including obesity, diabetes, hypertension, aging, chronic renal disease, and coronary artery disease, often accompanied by a significant pro-inflammatory state^5–7^. This biological and clinical heterogeneity is thought to contribute,^5,8^ at least in part, to the repeated neutral results in HFpEF randomized controlled trials. Unlike heart failure with reduced ejection fraction (HFrEF), which is frequently precipitated by a discrete index event such as acute myocardial infarction (MI), HFpEF more commonly reflects cumulative end-organ dysfunction arising from chronic, interacting comorbid conditions^9,10^.

A central challenge in HFpEF management is its marked clinical and biological heterogeneity, with distinct patient phenotypes associated with divergent disease trajectories and variable responses to treatment^5,11,12^. As a result, some patients experience clinically meaningful symptomatic improvement while others derive little or no benefit from the same interventions. In fact, unlike HFrEF, where patients benefit from guideline-directed medical therapies (GDMT)^13^, therapeutic options for HFpEF remain limited and mostly with variable effects^14^. This heterogeneity in treatment effect adds another layer of complexity which may contribute to the neutral or negative results of several large HFpEF trials.^15^

Another further methodological challenge lies in how “treatment response” is typically defined. Conventional responder analyses typically dichotomize patients as “responders” or “non-responders” based on whether they exceed a predefined threshold, such as ≥5-point improvement in the Kansas City Cardiomyopathy Questionnaire (KCCQ) overall summary score.^16^ Although thresholds can help us evaluate whether the degree of response is clinically important, dichotomizing continuous outcomes can limit both power and opportunities to uncover important signals.^17,18^

The proportion of “responders” observed in the treatment group versus the placebo group is not determined solely by the true treatment effect. Rather, it is highly sensitive to the underlying variability within each group, the choice of threshold, and the distributional properties of the outcome measure^18^. When the distributions of change scores in the treatment and placebo arms are similar in shape —reflected by approximately parallel cumulative distributions, the treatment effect is best summarized by the mean between-group difference. In such settings, there is no evidence of a distinct subgroup deriving qualitatively different benefit; instead, the appearance of a “responder “subgroup may arise as a consequence of arbitrary thresholding rather than true heterogeneity in treatment response.^18^ This limitation is particularly relevant in HFpEF where most clinical trials report modest or null mean treatment effects alongside substantial variability in patient-reported outcomes. In this context, a machine learning (ML) model trained to predict threshold-based “response” may achieve good discrimination while primarily capturing baseline disease severity, regression to the mean, and inherent natural variability in clinical trajectories, rather than true treatment heterogeneity in treatment effect. To address these limitations, analytic approaches that avoid dichotomization are required. Instead of modeling whether patients cross an arbitrary response threshold, the focus should shift to directly estimating treatment-by-variable interactions directly using continuous outcomes. This approach enables identification of baseline characteristics that modify the treatment effect itself, rather than merely capturing differences in underlying clinical trajectories.

In this study, we aim to address these limitations using two complementary analytic strategies designed to disentangle prognostic signals from treatment-specific effects. First, we develop a conventional ML responder model to identify baseline phenotypes associated with clinically meaningful improvement. We then formally evaluate whether this signal reflects treatment-specific benefit or underlying prognosis using permutation-based testing of treatment-by-subgroup interactions, demonstrating that the model predominantly captures prognostic variation.

Second, we implement an interaction-based individual treatment effect (ITE)^19^ model that explicitly encodes treatment-by-variable interactions using continuous outcomes. By jointly modeling treatment and placebo arms, this framework isolates baseline characteristics that differentially modify the treatment effect, thereby distinguishing drug-specific response from shared clinical trajectories.

We apply this framework across four HFpEF trials evaluating mechanistically distinct therapies: spironolactone (TOPCAT^20^), sildenafil (RELAX^21^), isosorbide mononitrate (NEAT-HFpEF^22^), and inorganic nitrite (INDIE-HFpEF^23^). We hypothesize that true treatment effect modifiers will be mechanism-specific, such that variables identified by the ITE models will demonstrate within-trial validity but will not generalize across therapies with distinct biological targets.

## 2. Methods

### a. Data Sources and Study Populations

The study utilized the HeartShare Extant Datasets Harmonized Clinical Trials Collection^24^ and was approved by the University of California, Davis Institutional Review Board (Protocol 2253505-2). From this harmonized repository, we selected four randomized clinical trials in HFpEF: TOPCAT, RELAX, NEAT-HFpEF, and INDIE-HFpEF. The Treatment of Preserved Cardiac Function Heart Failure with an Aldosterone Antagonist (TOPCAT) trial served as the primary training cohort^25^. TOPCAT randomized 3,445 patients with HFpEF (ejection fraction ≥ 45%) to spironolactone versus placebo. TOPCAT was selected as the training set due to its large sample size, comprehensive phenotypic characterization, well-defined clinical endpoints, and prior evidence of treatment effect heterogeneity, suggesting the presence of identifiable responder subgroups. For development of the prognostic responder model, analyses were restricted to participants assigned to the treatment arm.

We further included only individuals with both baseline and follow-up measurements for at least one clinically relevant outcome: Kansas City Cardiomyopathy Questionnaire (KCCQ) Overall Summary, New York Heart Association (NYHA) functional class, peak oxygen consumption (VO2 Max) or 6-minute walk test (6MWT). External validation was performed using three independent HFpEF trials, each evaluating different therapeutic interventions:

- RELAX: Phosphodiesterase-5 inhibition with sildenafil (n=216). This trial tested the hypothesis that phosphodiesterase 5 (PDE5) inhibition would improve exercise capacity through pulmonary vasodilation and enhanced ventricular-arterial coupling^26^.
- NEAT-HFpEF: Isosorbide mononitrate (organic nitrate) for exercise intolerance (n=110). This trial evaluated whether nitrate therapy would improve daily activity levels through preload reduction and enhanced exercise tolerance^27^.
- INDIE-HFpEF: Inorganic nitrite supplementation (n=105). This trial tested whether inorganic nitrite, converted to nitric oxide during exercise, would improve aerobic capacity through enhanced peripheral oxygen delivery^28^.

The diversity of therapeutic mechanisms across the validation cohorts provides a strict test of phenotype generalizability. In this context, consistent predictive performance across interventions would suggestthat the identified phenotypes reflect fundamental treatment-responsive biological phenotypes rather than mechanism-specific variables tied to a particular therapy.

### b. Outcome Definitions

#### Prognostic Model (Approach 1)

Treatment response was defined using a composite endpoint reflecting clinically meaningful improvement across multiple domains of HFpEF. Participants were classified as ‘responders’ if they achieved meaningful improvement in at least one of the following measures: NYHA class improvement ≥1 class^29^, KCCQ Overall Summary improvement ≥10 points^16^, or 6-minute walk distance improvement ≥45 meters^30^, or peak VO₂ improvement ≥1.5 mL/kg/min. This composite definition was selected to capturethe multidimensional and heterogeneous nature of clinical benefit in HFpEF. Also, we defined response as a KCCQ Overall Summary improvement of ≥10 points rather than the conventional ≥5-point threshold because, in preliminary analyses, the 5-point cutoff yielded unstable classification with substantial noise, likely reflecting measurement variability at the minimal clinically important difference (MCID); the 10-point threshold produced more stable and clinically interpretable responder definitions.

#### ITE Model (Approach 2)

The primary outcome was defined as the continuous change in score (ΔKCCQ, Δ6MWT, or ΔVO₂ depending on the training trial) from baseline to follow-up. The use of continuous outcomes (i.e., excluding the NYHA class), rather than threshold-based definitions, preserves the full distribution of treatment response and enables direct estimation of treatment-by-variable interactions within a regression framework.

### c. Data Preprocessing

All models were developed using baseline variables extracted from the HeartShare harmonized dataset and the analisys was performed on the NHLBI BioData Catalyst ecosystem.^31^ To ensure data quality and reliable imputation, variables with more than 30% missingness or not available across all studies were excluded, balancing retention of clinically relevant variables with robustness of imputation. A full list of the baseline variables used in presented in **Table 1**.

**Table 1.**
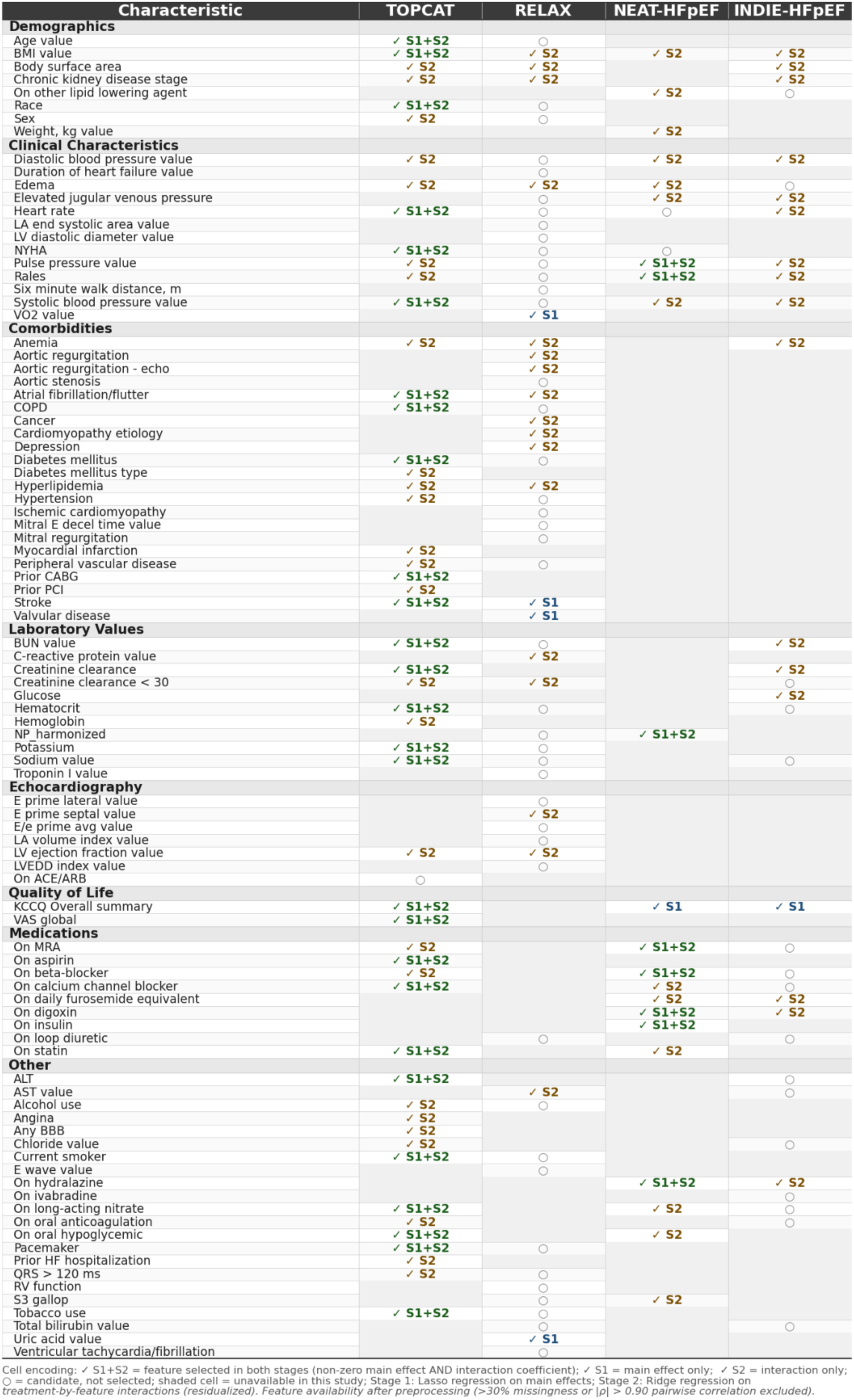
Baseline variable inventory and selection status across the four ITE models. Each row represents a candidate baseline variable, grouped by clinical category; each column represents one trial-specific interaction-based ITE model (TOPCAT, RELAX, NEAT-HFpEF, INDIE-HFpEF). Cell encoding: ✓ S1+S2 = non-zero coefficient in both Stage 1 (Lasso main effect) and Stage 2 (Ridge treatment-by-variable interaction); ✓ S1 = selected as main effect only; ✓ S2 = selected as treatment-modifying interaction only; ○ = candidate variable available to the model but not selected (coefficient shrunk to zero); shaded cell = variable unavailable in that trial after preprocessing (>30% missingness or |ρ| > 0.90 pairwise correlation with another variable). This table demonstrates that each trial’s interaction-based model selected a distinct, drug-specific subset of treatment-modifying variables from a largely overlapping candidate pool, supporting the specificity of the identified phenotype–treatment associations rather than a shared prognostic signal.

Missing data were imputed Singular Value Decomposition (SVD)^32^, chosen over simpler methods (mean/median imputation) for several reasons, including preserving correlation structure among variables, which is critical when variables are biologically related (e.g., creatinine and eGFR, BNP and NYHA class). SVD also naturally handles the multicollinearity common in clinical data, and iterative refinement converges to stable estimates that respect the underlying data manifold.

The SVD imputer was configured with 5 components, 10 maximum iterations, and a convergence tolerance of 1e-5. Internal standardization was applied before SVD to prevent variables with larger scales from dominating the decomposition.

Highly correlated variable pairs (Pearson r > 0.6) were identified, and one member of each pair was removed to reduce multicollinearity. When removing correlated variables, preference was given to retaining variables with lower missingness rates, established clinical interpretability, and those available across all validation cohorts. Variables were standardized to a zero mean and unit variance using parameters derived solely from the training dataset, which were then applied to validation sets to prevent information leakage.

### d. Model Development

#### Approach 1: Prognostic Responder Classifier

To evaluate whether conventional responder modeling predominantly captures prognostic rather than treatment-specific signals, we developed a regularized classification model to predict clinically meaningful improvement. Specifically, an L1-regularized (LASSO) logistic regression model was trained using participants assigned to the treatment arm of TOPCAT (n = 877).

Feature selection was performed using recursive variable elimination with cross-validation (RFECV). To minimize overfitting and ensure unbiased performance estimation, model development and evaluation were conducted within a nested 5-fold cross-validation framework, maintaining strict separation between feature selection and model assessment. Additional details regarding model specification and tuning are provided in the Supplementary Methods.

#### Approach 2: Two-Stage Interaction-Based ITE Model

To distinguish treatment-specific effects from underlying prognostic variation, we developed a two-stage interaction-based modeling framework. This approach was designed to separate baseline characteristics that modify the causal effect of treatment from those that are associated with overall clinical trajectories independent of treatment assignment.

Leveraging data from both treatment and placebo arms, the model was structured in two sequential stages, each targeting a distinct source of outcome variability. The first stage accounts for prognostic variation shared across treatment groups, while the second stage estimates treatment-by-variable interactions to isolate characteristics that differentially modify the treatment effect.

#### Data Preprocessing

All preprocessing steps were conducted separately within each analytical unit (training set, cross-validation fold, or external validation cohort) to ensure strict separation between model development and evaluation. To prevent information leakage, imputation and collinearity assessments were performed exclusively on the training partition and subsequently applied to held-out data. All variables were standardized to zero mean and unit variance using parameters derived from the training data.

For trials with smaller sample sizes (N < 300; RELAX, NEAT-HFpEF, and INDIE-HFpEF), an adaptive variable pre-selection step was implemented prior to construction of interaction terms to mitigate overfitting. The maximum number of interaction variables was constrained to N/10 (minimum of 5), consistent with the heuristic of maintaining at least 10 observations per variable for stable estimation.

Candidate variables for interaction terms were ranked using a univariate interaction-signal proxy. Specifically, for each variable *X_j_*, we computed the absolute Spearman correlation between the interaction term *X_j_*×*T* and the outcome (*Y*), and penalized this score by subtracting one-half of the absolute marginal correlation between *X_j_* and *Y*. This scoring approach prioritizes variables that preferentially modify treatment effects over those that primarily reflect prognostic associations. Only the top-ranked variables were selected for inclusion as interaction terms, while all variables were retained as main effects in Stage 1. For TOPCAT (N = 1,536), this pre-selection step was not applied, as the larger sample size permitted inclusion of interaction terms for all candidate variables without compromising model stability.

#### Stage 1: Capturing Prognostic Effects

In the first stage, we built a prognostic model to identify baseline variables associated with outcomes, regardless of whether a patient received treatment or a placebo. The model takes the form:

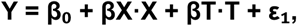

where Y is the continuous change score (for example, change in KCCQ from baseline to last follow-up), X represents baseline variables, T is a binary treatment indicator (1 = treatment, 0 = placebo), βX captures prognostic effects (i.e., variables that predict outcome in general) and βT captures the average treatment effect across the population.

We estimated this model using L1-regularized linear regression (Lasso).In practice, this means that only the strongest prognostic variables should remain in the model. The regularization parameter was chosen using 5-fold cross-validation. After fitting Stage 1, we computed as follows:

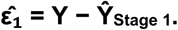

These residuals represent what’s *left over* (i.e., the part of outcome variation not explained by prognostic baseline variables or the average treatment effect). This can be considered the part of the signal that accounts for the remaining effects of treatment-specific heterogeneity.

#### Stage 2: Identifying Treatment-Modifying Interactions

In the second stage, we modeled those residuals from Stage 1 as a function of treatment–variable interaction terms:

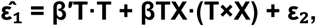

where T×X captures whether the relationship between a variable and the outcome differs between treatment and placebo arms. By design, these coefficients (βTX) can only be non-zero if a variable modifies the treatment effect.

For this stage, we used L2-regularized linear regression (Ridge regression). In the cases where Ridge produced practically no interaction signal (maximum absolute coefficient < 10⁻⁶), we switched to Elastic Net with low L1 ratios (0.1–0.5). Both the L1 ratio and regularization strength were selected by cross-validation.

In summary, by separating the steps (i.e., first modeling prognostic effects, then modeling interactions on the residuals), we ensure that Stage 2 focuses specifically on treatment-related variation that remains after accounting for baseline prognosis. In a single regularized model, strong prognostic effects (i.e., variables that predict outcome regardless of treatment) dominate the penalty structure and shrink the weaker treatment-modifying interactions toward zero. By removing prognostic variation in Stage 1, the Stage 2 residuals isolate specifically who benefits more or less from the active treatment, allowing interaction terms to be estimated without competing against stronger main effects. This characteristic is particularly important for the smaller trials where the number of variables can quickly outpace the number of observations and subtler interaction terms are dominated by main effects.

#### Estimation of Individual Treatment Effect (ITE)

We computed the individual treatment effect (ITE) for patient *i* using the Stage 2 interaction coefficients:

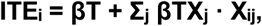

where βT represents the combined treatment coefficient from both stages, and βTXⱼ are the interaction coefficients from Stage 2. We define participants with higher predicted ITE values as those whose baseline characteristics suggest greater treatment benefit.

We trained separate ITE models for each trial with both arms available, using each trial’s own primary endpoint as the outcome: ΔKCCQ for TOPCAT, NEAT-HFpEF, and INDIE-HFpEF, while Δ6MWT for RELAX. We chose continuous outcome measures because more effectively capture treatment-modifying effects in interaction models.

#### Model Performance Assessment

To obtain performance estimates of the proposed 2-stage model, we performed 5-fold cross-validation. Importantly, the *entire pipeline* was repeated within each fold and not just the model fitting. For each fold, the training data were preprocessed independently (median imputation, standardization, and collinearity filtering computed on training data only). Next, the Stage 1 Lasso model is trained, and residuals are computed from Stage 1 predictions. The Stage 2 model is then trained on those residuals. Predictions for the held-out fold were generated by combining outputs from both stages, using preprocessing parameters from the training partition. Finally, cross-validated R² and RMSE were calculated from the pooled held-out predictions across all folds. We adopted this design to ensure that no information from the test folds influences preprocessing or model training (i.e., minimizing the risk of information leakage when estimating treatment heterogeneity).

#### Model Validation

##### i. Permutation-Based Interaction Test for the Prognostic Model (Stage 1)

To test whether the prognostic classifier described above captures treatment-specific signal, we applied it to both treatment and placebo arms. To do so, for each study, we applied the model to both treatment and placebo arms, and, based on their variables, patients were classified as predicted responders or non-responders. Next, we computed treatment effects within each subgroup. If both subgroups show similar treatment–placebo differences (interaction ≈ 0), then the model is purely prognostic. A positive interaction statistic indicates that the model identifies patients who differentially benefit from the active treatment beyond what baseline prognosis alone would predict, confirming that the ITE captures a genuinely prescriptive rather than merely prognostic signal.

##### ii. Complete ITE Model Validation

The central validation aims to test whether patients predicted to have high ITE show larger treatment effects than those predicted to have low ITE. To assess if this is true, we first split trial participants at the median predicted ITE in the treatment arm. Next, the same threshold is applied to the placebo arm, and we compute the treatment effect (treatment mean minus placebo mean) separately in the high-ITE and low-ITE groups. The interaction contrast was defined as:

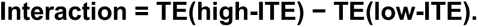

If the Interaction value is positive, it means patients predicted to benefit more actually show a larger treatment–placebo difference. We perform a permutation test with 1,000 permutations, shuffling treatment assignments while keeping the ITE subgroup labels fixed, to assess the statistical significance of the results. This is done to preserve the model’s stratification while breaking any true treatment signal, and should produce a null distribution.

We also perform a drug specificity validation, where for each ITE trial model, we test if it generalizes to trials using drugs with a similar mechanism, or if it only works in its own trial. This first test aims to identify treatment-modifying variables that reflect pharmacologically meaningful changes. The second test could indicate overfitting or trial-specific confounding.

We also calculated Spearman rank correlations among predicted ITE and the observed outcome change within the treatment arm of each validation cohort. A significant positive correlation in the training trial provides internal validation. Correlations in external trials test whether the treatment-modifying signal is portable.

### f. Surrogate Decision Tree for Clinical Interpretability

Although the Ridge interaction model produces a continuous ranking of treatment-modifying variables, its linear combination of dozens of coefficients is difficult to translate into actionable clinical decision rules. To make the results more interpretable and actionable, we built a surrogate decision tree for each trial. The idea is not to replace the proposed interaction model, but to approximate it in a way that produces clear, rule-based decisions.

For each study, we first computed the predicted ITE for every patient in the training set using the full two-stage model. Then we trained a decision tree regressor (maximum depth = 3, so at most 8 terminal nodes) to predict those ITE scores using the baseline variables. Importantly, we purposely used the unstandardized variables so that splits in the decision tree appear in clinically meaningful units (e.g., “BUN ≤ 37.5 mg/dL”) rather than abstract z-scores. To avoid unstable or overly specific splits, we required each leaf to contain at least 5% of the sample (i.e., at least 10 patients). **Figure 1** describes the phases and validation steps for the proposed ITE model.

**Figure 1.**
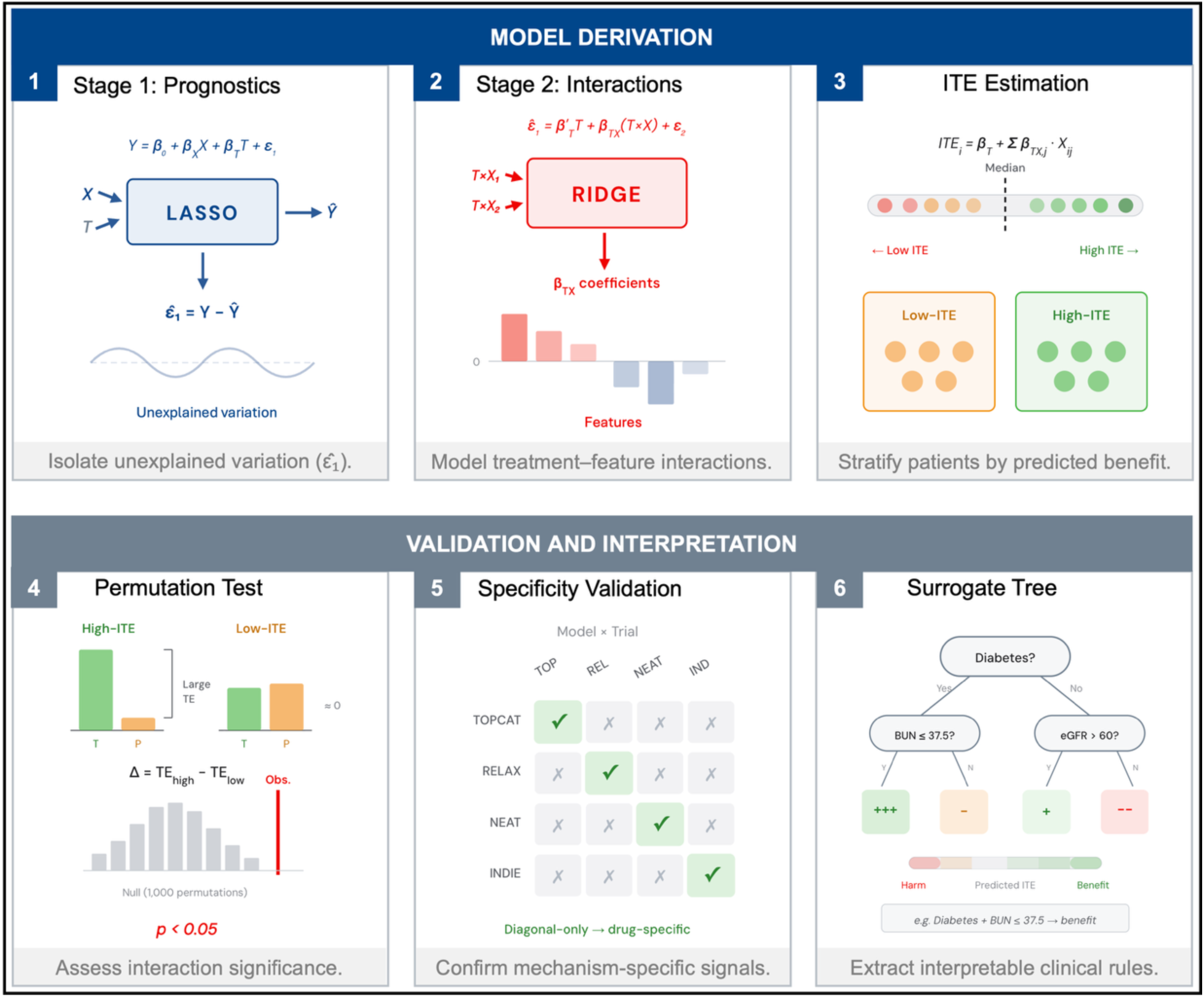
Representation of the proposed model and validation pipeline. Stage 1 uses Lasso regression (a method that selects the most relevant predictors by shrinking less important ones to zero) to remove the effect of baseline disease severity on outcomes, so that Stage 2 can use Ridge regression (a method that retains all candidate predictors while preventing any single variable from dominating) to isolate which patient variables specifically modify treatment benefit. Each patient receives a predicted treatment effect score (ITE) and is classified as higher or lower predicted benefit. The model is validated through permutation testing to confirm that the subgroup separation is statistically significant, cross-trial analysis to confirm that the identified variables are specific to each drug’s mechanism of action, and surrogate decision trees to translate the model into clinically interpretable rules using routinely available variables.

## 3. Results

### a. Study Population and Baseline Characteristics

For the prognostic model, 701 participants from the treatment arm of TOPCAT with available baseline and follow-up functional data were included, and 180 (25.7%) met the composite responder definition. Baseline characteristics are shown in **Table 2**. Responders were slightly younger (70.4 ± 10.2 vs. 72.4 ± 9.5 years, p=0.022), had a higher body mass index, and had worse baseline KCCQ, visual analog scale (VAS) for global health, and NYHA class, indicating a greater initial symptom burden. They also presented had higher SBP and DBP and less aspirin users No statistically significant difference in sex, race, ejection fraction, or most other medical conditions was identified. For the ITE model, we pooled the full TOPCAT cohort, comprising 1,536 patients (783 in the treatment arm and 753 in the placebo arm). We also performed external validation on RELAX (n=191; 94 treatment, 97 placebo), NEAT-HFpEF (n=215; 108 treatment, 107 placebo), and INDIE-HFpEF (n=196; 98 treatment, 98 placebo).

**Table 2.** Baseline characteristics of responders and non-responders in the TOPCAT treatment arm (n=701). The responders were slightly younger (around 70 vs. 72 years old and had higher BMI. They also present worse baseline values of KCCQ, VAS Scale, and NYHA. This group can be characterized by greater initial symptom burden. Values are mean ± SD, median (IQR), or n (%). P values: Mann-Whitney U or Chi-square/Fisher. SMD = standardized mean difference. Values are mean ± SD, median (IQR), or n (%). P values: Mann-Whitney U or Chi-square/Fisher. SMD = standardized mean difference.

| Characteristic | Non-Responders (n=521) | Responders (n=180) | P Value | SMD |
| --- | --- | --- | --- | --- |
| **Demographics** |  |  |  |  |
| Age, years | 72.4 ± 9.5 | 70.4 ± 10.2 | **0.022** | 0.21 |
| Body mass index, kg/m<sup>2</sup> | 33.4 ± 7.6 | 35.2 ± 8.8 | **0.022** | 0.23 |
| Body surface area, m<sup>2</sup> | 2.0 ± 0.3 | 2.0 ± 0.3 | 0.733 | 0.00 |
| Female sex | 0 (0.0) | 0 (0.0) | 1.000 |  |
| Race |  |  | 0.074 |  |
| 1.0 | 357 (68.5) | 106 (58.9) |  |  |
| 2.0 | 66 (12.7) | 35 (19.4) |  |  |
| 3.0 | 88 (16.9) | 36 (20.0) |  |  |
| 6.0 | 10 (1.9) | 3 (1.7) |  |  |
| **Clinical Characteristics** |  |  |  |  |
| Systolic BP, mm Hg | 126.1 ± 15.8 | 128.9 ± 13.5 | **0.027** | 0.19 |
| Diastolic BP, mm Hg | 70.8 ± 11.0 | 73.0 ± 12.2 | **0.042** | 0.19 |
| Pulse pressure, mm Hg | 55.3 ± 14.3 | 55.9 ± 12.6 | 0.341 | 0.05 |
| NYHA |  |  | **<0.001** |  |
| 2.0 | 397 (76.3) | 56 (31.1) |  |  |
| 3.0 | 123 (23.7) | 124 (68.9) |  |  |
| **Comorbidities** |  |  |  |  |
| Diabetes mellitus | 223 (42.8) | 81 (45.0) | 0.670 | 0.04 |
| Atrial fibrillation | 226 (43.4) | 68 (37.8) | 0.221 | 0.11 |
| Prior MI | 98 (18.8) | 39 (21.7) | 0.469 | 0.07 |
| Prior CABG | 102 (19.6) | 26 (14.4) | 0.154 | 0.13 |
| COPD | 98 (18.8) | 29 (16.1) | 0.485 | 0.07 |
| CKD | 195 (37.4) | 62 (34.4) | 0.531 | 0.06 |
| Prior stroke | 41 (7.9) | 13 (7.2) | 0.906 | 0.02 |
| PVD | 77 (14.8) | 20 (11.1) | 0.270 | 0.11 |
| **Laboratory Values** |  |  |  |  |
| Hemoglobin, g/dL | 12.9 ± 1.6 | 12.7 ± 1.9 | 0.131 | 0.09 |
| Hematocrit, % | 46.1 ± 169.0 | 38.3 ± 5.1 | 0.176 | 0.05 |
| Sodium, mEq/L | 139.9 ± 3.1 | 139.4 ± 3.4 | 0.160 | 0.15 |
| Potassium, mEq/L | 4.2 ± 0.4 | 4.1 ± 0.4 | 0.103 | 0.14 |
| BUN, mg/dL | 25.2 ± 13.0 | 23.8 ± 11.3 | 0.227 | 0.11 |
| CrCl, mL/min | 60.9 ± 19.8 | 61.3 ± 20.7 | 0.981 | 0.02 |
| Glucose, mg/dL | 110.3 ± 41.6 | 119.1 ± 57.1 | 0.599 | 0.19 |
| NT-proBNP/BNP (harmonized) | 1656.0 (960.0-3216.0) | 1362.0 (858.0-2790.0) | 0.125 | 0.22 |
| **Echocardiography** |  |  |  |  |
| LVEF, % | 58.1 ± 7.4 | 58.5 ± 7.8 | 0.682 | 0.05 |
| LV mass index, g/m<sup>2</sup> | 106.2 ± 27.0 | 113.4 ± 34.1 | 0.339 | 0.25 |
| E/e' ratio | 14.4 ± 7.0 | 13.3 ± 5.3 | 0.754 | 0.17 |
| RVSP, mm Hg | 41.1 ± 12.8 | 38.4 ± 11.2 | 0.399 | 0.22 |
| **Quality of Life** |  |  |  |  |
| KCCQ Overall Summary | 61.7 ± 23.3 | 47.4 ± 22.0 | **<0.001** | 0.62 |
| VAS global health | 64.7 ± 19.6 | 58.9 ± 21.1 | **<0.001** | 0.29 |
| **Medications** |  |  |  |  |
| ACEI/ARB | 0 (0.0) | 0 (0.0) | 1.000 |  |
| Beta-blocker | 111 (21.3) | 46 (25.6) | 0.282 | 0.10 |
| CCB | 338 (64.9) | 105 (58.3) | 0.139 | 0.14 |
| MRA | 0 (0.0) | 0 (0.0) | 1.000 |  |
| Loop diuretic | 0 (0.0) | 0 (0.0) | 1.000 |  |
| Statin | 183 (35.1) | 67 (37.2) | 0.677 | 0.04 |
| Aspirin | 208 (39.9) | 89 (49.4) | **0.032** | 0.19 |
| Anticoagulant | 340 (65.3) | 123 (68.3) | 0.510 | 0.06 |

### b. Approach 1: Prognostic Responder Classifier

#### Internal Validation

The model results suggest robust prediction of response rate probabilities across the risk spectrum in internal validation. The model achieved an internal AUROC of 0.76 (95% CI: 0.68–0.84) with AUPRC of 0.50 (**Figure 2A**). Calibration results also indicate strong performance with a slope of 0.899. Using the SHapley Additive exPlanations (SHAP) framework^33^ to interpret the model’s decision identified baseline NYHA functional class and KCCQ score as the dominant predictors (**Figure 2B**). A higher NYHA class was strongly associated with response probability (mean |SHAP| = 0.19); patients with Class III symptoms had substantially higher predicted response rates than those with Class II symptoms. Similarly, lower baseline KCCQ scores predicted greater improvement (i.e., patients with scores below 50 were markedly more likely to respond than those with higher baseline quality-of-life scores).

**Figure 2.**
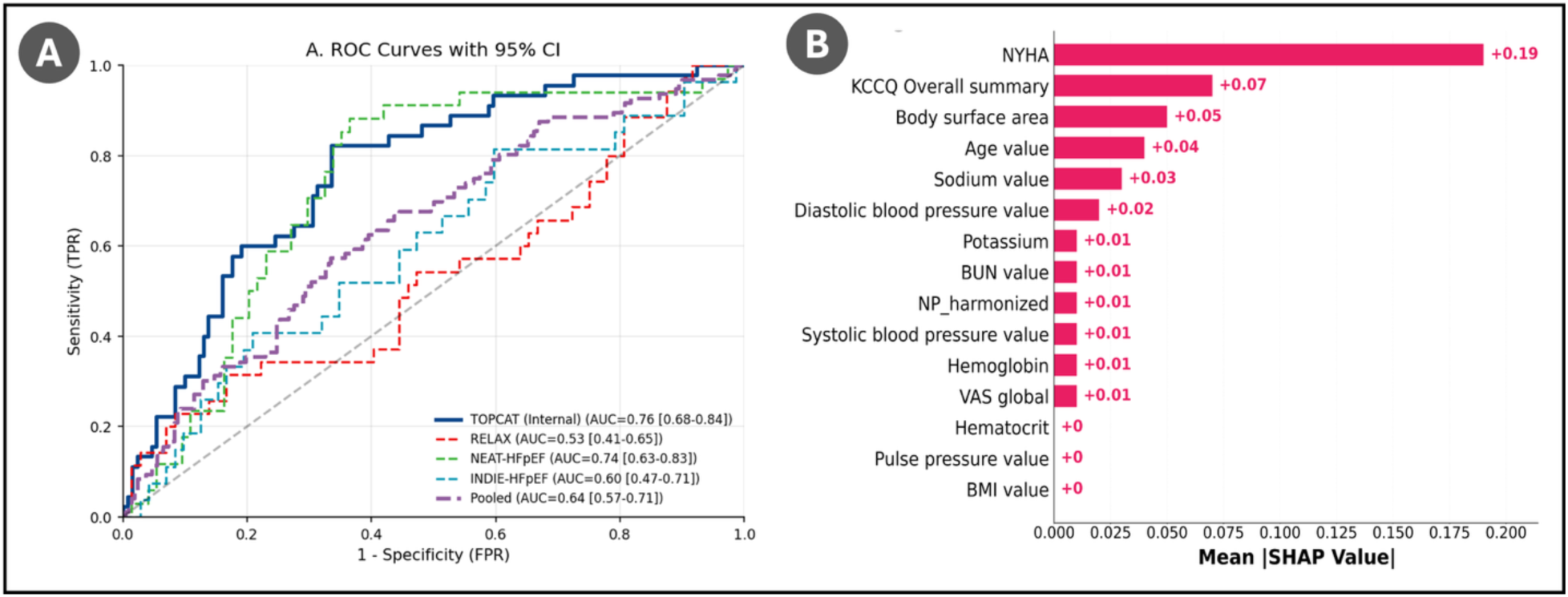
Prognostic model discrimination and variable importance for predicting clinical deterioration in HFpEF. (A) Receiver operating characteristic (ROC) curves with 95% confidence intervals for the TOPCAT-trained prognostic model evaluated across four HFpEF clinical trials and a pooled external validation cohort. The model demonstrated satisfactory internal discrimination in TOPCAT (AUC = 0.76) and external generalizability to NEAT-HFpEF (AUC = 0.74), with more modest/poor performance in INDIE-HFpEF (AUC = 0.60) and RELAX (AUC = 0.53). (B) Mean absolute SHAP values indicating the relative contribution of each baseline covariate to model predictions. NYHA functional class and KCCQ Overall Summary Score were the dominant predictors.

#### External Validation

In the external validation (**Figure 2A**), the model generalized best to NEAT-HFpEF (isosorbide mononitrate; AUC 0.74 [95% CI: 0.63–0.83]), with sensitivity of 79.4% and specificity of 66.2%. Performance in INDIE-HFpEF (inorganic nitrite) and the pooled external cohort was modest (AUC 0.60 [95% CI: 0.47–0.71]) and (AUC 0.53 [95% CI: 0.41–0.65]), respectively. Lastly, performance on RELAX was near chance with an AUC of 0.53 [95% CI: 0.41–0.65]. The result on RELAX could be linked to the different primary endpoint (6MWT vs. KCCQ) used for training and the distinct population characteristics between the trials.

#### Prognostic Value: Survival Analysis

We also assessed the prognostic evaluation for participants in the responder and non-responder groups based on TOPCAT data. Using a set of five indicators, we assessed long-term cardiovascular outcomes over a median follow-up of 3.3 years. **Figure 3** and **Table 3** shows that non-responders experienced higher rates of cardiovascular death or heart failure hospitalization (33.6% vs. 21.5%; hazard ratio [HR] 1.77; 95% CI: 1.33–2.36; p<0.001). HF hospitalization (HR 1.92; 95% CI: 1.38–2.68; p<0.001) was the primary driver of this association, with trends toward increased all-cause mortality (HR 1.42; 95% CI: 1.02–1.96; p=0.035) and cardiovascular mortality (HR 1.56; 95% CI: 0.99–2.46; p=0.052). Kaplan-Meier curves diverged within 6 months and maintained separation throughout follow-up. These findings suggest that prognostic phenotypes identified present differential long-term risk which could also be associated with modifiable disease trajectories of the responder group.

**Figure 3.**
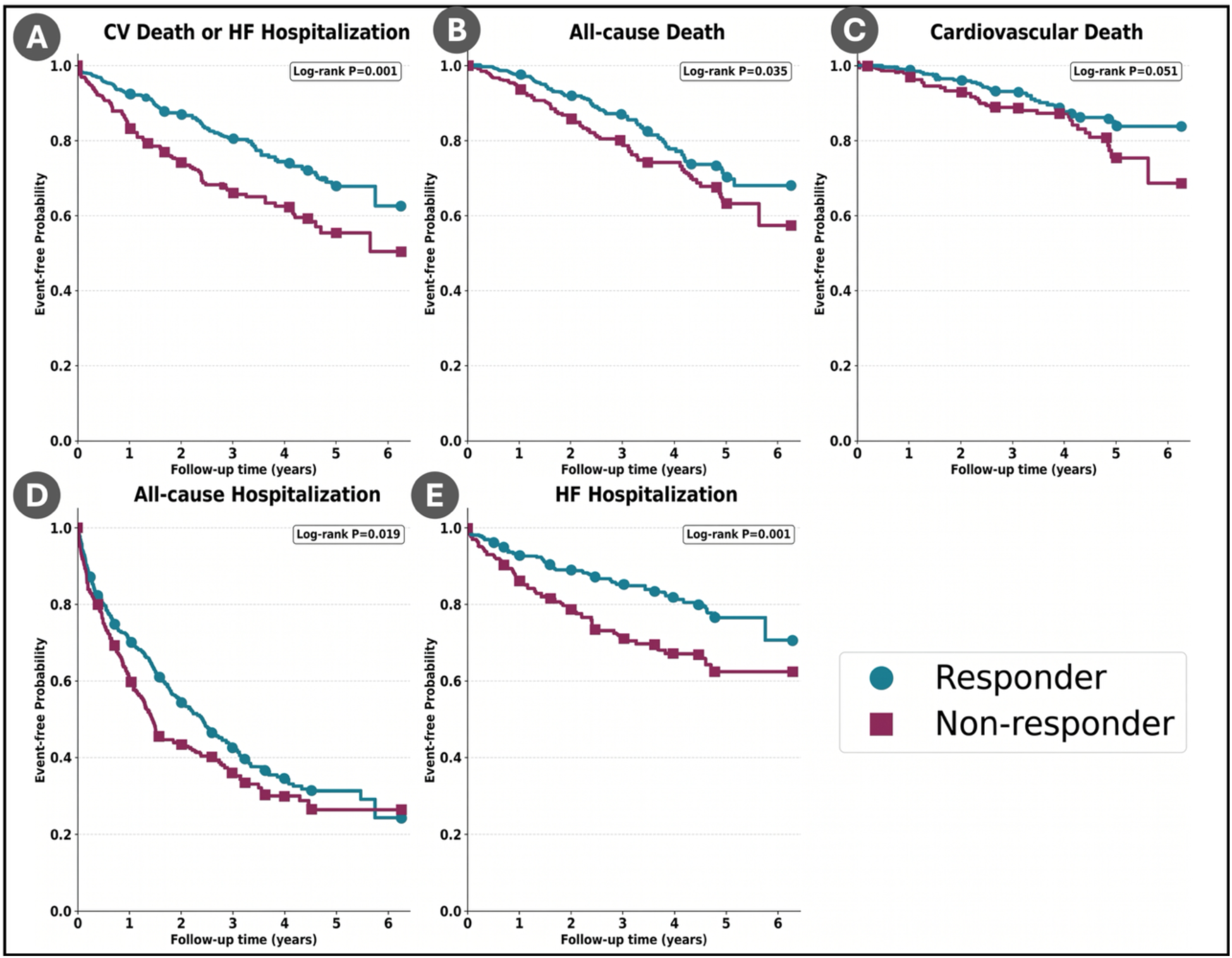
Kaplan-Meier event-free survival curves stratified by predicted treatment response in TOPCAT. Patients classified as predicted responders (blue) versus non-responders (purple) to spironolactone were compared across five clinical endpoints: (A) the primary composite of cardiovascular death or heart failure hospitalization, (B) all-cause death, (C) cardiovascular death, (D) all-cause hospitalization, and (E) heart failure hospitalization. Log-rank p-values are shown for each comparison. Predicted responders demonstrated significantly better event-free survival for the primary composite endpoint (P < 0.001) and heart failure hospitalization (P < 0.001).

**Table 3.** Summary of the clinical outcomes for Responders (n=424) and Non-responders (n=277). The upper section details the frequency of events (n, %) for categories such as cardiovascular (CV) death, heart failure (HF) hospitalization, and all-cause mortality, with significance tested using the log-rank test. The lower section presents Unadjusted Hazard Ratios (HR) with 95% Confidence Intervals (CI), utilizing Responders as the reference group (1.0) and calculating significance via Cox proportional hazards regression (Wald test). * P<0.05; † P≤0.01; ‡ P≤0.001.

| Outcome, n (%) | Responder (n=424) | Non-responder (n=277) | Log-rank P |
| --- | --- | --- | --- |
| CV Death or HF Hospitalization | 91 (21.5%) | 93 (33.6%) | <0.001 |
| All-cause Death | 77 (18.2%) | 68 (24.5%) | 0.035 |
| Cardiovascular Death | 38 (9.0%) | 37 (13.4%) | 0.051 |
| All-cause Hospitalization | 244 (57.5%) | 176 (63.5%) | 0.019 |
| HF Hospitalization | 67 (15.8%) | 75 (27.1%) | <0.001 |
| Unadjusted HR (95% CI) |  |  | Cox P |
| CV Death or HF Hospitalization | 1.0 (Ref) | 1.77 (1.33-2.36)‡ | <0.001 |
| All-cause Death | 1.0 (Ref) | 1.42 (1.02-1.96)* | 0.036 |
| Cardiovascular Death | 1.0 (Ref) | 1.56 (0.99-2.46) | 0.053 |
| All-cause Hospitalization | 1.0 (Ref) | 1.26 (1.04-1.53)* | 0.019 |
| HF Hospitalization | 1.0 (Ref) | 1.92 (1.38-2.68)‡ | <0.001 |

Reference Group: Responder. \* P<0.05; † P≤0.01; ‡ P≤0.001.
P values for event counts from log-rank test; P values for HR from Cox proportional hazards regression (Wald test).

#### Indication of Prognostic Rather Than Predictive Signal

The model performs well on endpoints and survival outcomes and generalizes well across the HFpEF trials assessed. This combination suggests that the model is not identifying people who specifically respond to treatment but rather recognizing those patients with inherently better or worse prognosis. If this were truly a predictive model of treatment response, we would expect the “responders” to benefit more from spironolactone than from placebo. However, as shown in **Figure 4**, this was not the case. This is also supported by ITE model results (see Approach 2 below), in which the predicted subgroups from the prognostic model were tested for a treatment-by-subgroup interaction with spironolactone assignment resulting in a nonsignificant interaction (p = 0.17). In other words, there was no evidence that the treatment effect differed between the predicted groups.

**Figure 4.**
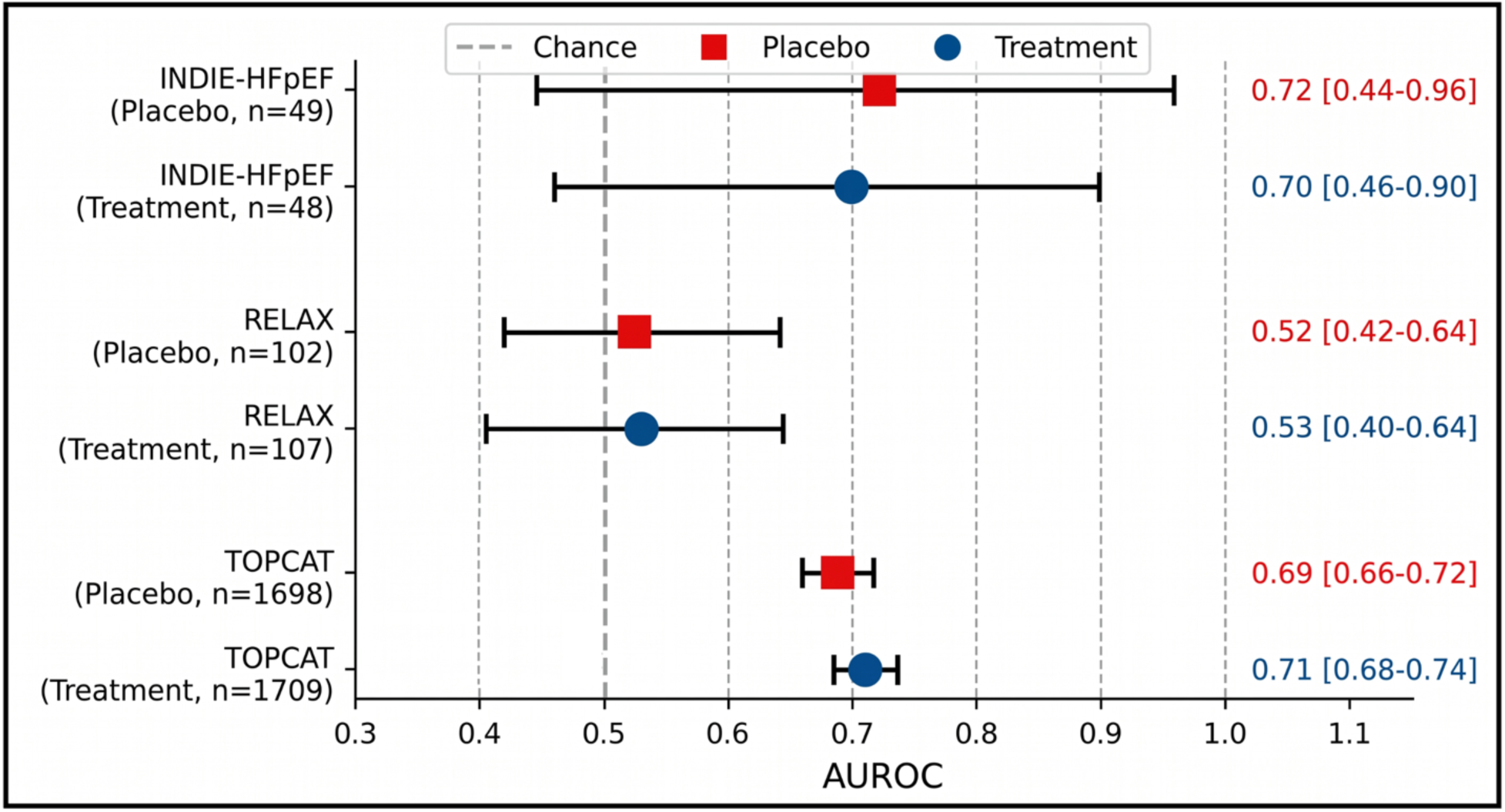
Prognostic model discrimination by treatment arm across HFpEF clinical trials. Area under the receiver operating characteristic curve (AUROC) with 95% confidence intervals for the prognostic model applied separately to treatment and placebo arms in TOPCAT, RELAX, and INDIE-HFpEF. Comparable AUROC values between treatment and placebo arms within each trial indicate that model predictions reflect baseline prognostic risk rather than treatment-specific effects. The dashed line indicates chance-level discrimination (AUROC = 0.5).

### c. Approach 2: Two-Stage Interaction-Based ITE Model

#### Two-Stage Model Architecture

We trained separate ITE models for each HFpEF trial, and for TOPCAT, the Lasso model (Stage 1) explained a significant portion of the outcome variance (R²=0.298). For the smaller trials, a moderate variance was observed (RELAX: R²=0.237; NEAT-HFpEF: R²=0.152; INDIE-HFpEF: R²=0.100). Interestingly, the treatment coefficient (βT) was near zero in Stage 1 across all trials, consistent with the overall null or modestly positive results reported by these studies. The results of Stage 2 (i.e., Ridge regression on residuals), on the other hand, suggest that it captured the treatment-modifying interaction terms, with full-model cross-validated R² values of 0.266 (TOPCAT), 0.070 (RELAX), 0.058 (NEAT-HFpEF), and 0.068 (INDIE-HFpEF).

#### TOPCAT (Spironolactone): Treatment-Modifying Variables

The ITE model for TOPCAT identified five main variables that modify treatment response. As shown in **Figure 5A**, all the listed variables can be considered variables in cardiorenal patients. Some appear to promote increased benefit from spironolactone while some appear to be linked with reduced benefit. For example, creatinine clearance below 30 mL/min (βTX=+0.110) and the presence of any bundle branch block (βTX=+0.064) were associated with increased benefit from spironolactone. In contrast, diabetes mellitus (βTX=−0.108), use of oral hypoglycemic agents (βTX=−0.100), and elevated BUN (βTX=−0.084) were linked to reduced benefit. Additional modifiers included statin use, aspirin use, prior coronary artery bypass grafting (CABG), QRS duration greater than 120 ms, and atrial fibrillation or flutter. Together, these variables suggest a cardiorenal phenotype as the principal determinant of spironolactone response in HFpEF. This result is supported by post hoc TOPCAT analyses, who identified an obese with chronic kidney disease and elevated renin as the subgroup most responsive to spironolactone^34^, while showing the potential adverse effects of spironolactone for a diabetic phenotype^35,36^. The baseline characteristics of patients in the treatment arm stratified by predicted ITE are described in **Table 4**.

**Figure 5.**
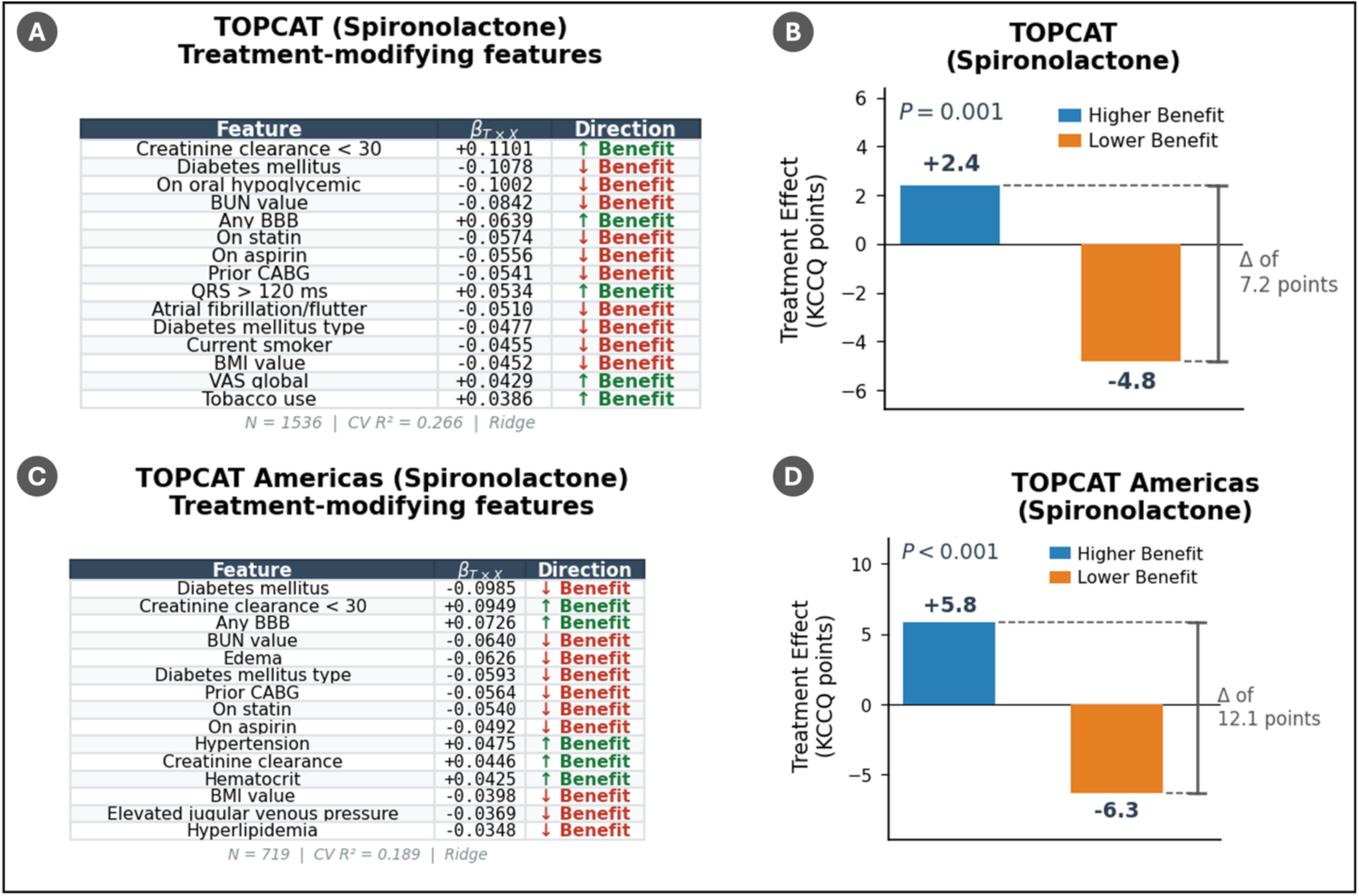
Interaction-based treatment effect model trained on TOPCAT (Spironolactone), comparing the full cohort with the Americas-only cohort. (A, C) Non-zero treatment–covariate interaction coefficients (β_T×X) from the penalized Ridge regression model, ranked by absolute magnitude. Positive coefficients (green) indicate variables associated with greater predicted treatment benefit, while negative coefficients (red) indicate variables associated with reduced benefit or predicted harm. Panel A shows the full TOPCAT cohort (N = 1,536; CV R² = 0.266) and Panel C shows the Americas-only cohort after exclusion of Russia/Georgia sites (N = 719; CV R² = 0.189). Core treatment-modifying variables (i.e., diabetes mellitus, creatinine clearance < 30, BUN value, and bundle branch block) are identified in both cohorts, though their relative ranking shifts with the restricted sample. (B, D) Differential treatment effect stratified by predicted individualized treatment effect (ITE). Patients predicted to have higher benefit (blue) versus lower benefit (orange) were compared based on the difference in outcome change between treatment and placebo arms (TE = Δ_Tx − Δ_Pb). Restricting to the Americas cohort (D) amplified the treatment effect separation from 7.2 (B) to 12.1 KCCQ points (both P ≤ 0.001), consistent with stronger treatment-response heterogeneity once the Russia/Georgia enrollment concerns are addressed.

**Table 4.**
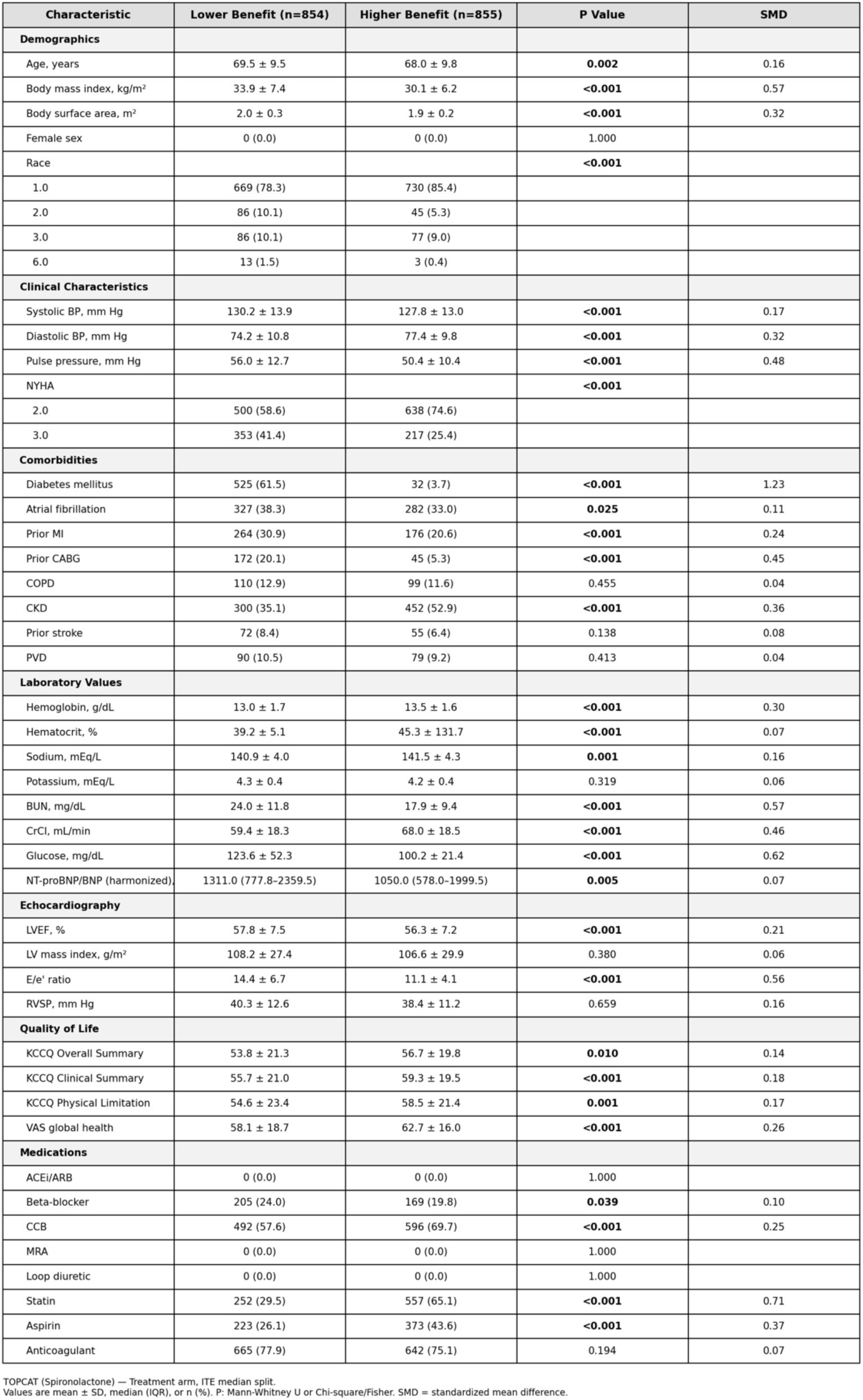
Baseline characteristics of TOPCAT (Spironolactone) patients in the treatment arm stratified by predicted ITE median split. Patients classified as Higher Benefit (n = 855) versus Lower Benefit (n = 854) differed significantly across multiple domains, including demographics, comorbidity burden, renal function, and medication use. The Lower Benefit subgroup was characterized by higher BMI, greater prevalence of diabetes mellitus (61.5% vs. 3.7%), elevated BUN and glucose, lower creatinine clearance, higher E/e′ ratio, and more advanced NYHA class which is consistent with the cardiorenal and metabolic phenotype identified by the interaction model as predictive of reduced spironolactone response.

Application of the model to the TOPCAT population revealed substantial heterogeneity in treatment effects. **Figure 5B** shows that patients with predicted high ITE experienced a treatment benefit of +2.4 KCCQ points (treatment: +11.3, placebo: +8.9). In contrast, those with predicted low ITE exhibited a treatment effect of −4.8 KCCQ points (treatment: +4.6, placebo: +9.4), resulting in an interaction statistic of +7.2 (permutation p=0.001).

These findings suggest that the overall null result in TOPCAT may mask significant heterogeneity, with some patients (i.e., those with a high predicted ITE) seeming to benefit from spironolactone as they present clinically meaningful improvements not observed in a similar placebo group. On the other hand, patients with a low predicted ITE may not only fail to benefit but also be harmed by the treatment compared to the placebo group. In this scenario, rather than a neutral classification of the trial, the reality seems more nuanced: spironolactone may help certain patients while potentially disadvantaging others, which is a very important finding for personalized treatment decisions in HFpEF.

A known limitation of the TOPCAT trial concerns the recruitment of patients who may not have had true HFpEF. Post hoc analyses identified patients’ enrollment fidelity and endpoint adjudication at Russia/Georgia sites^37–40^. To address these concerns, we repeated the ITE analysis, restricting it to the American cohort. Of 3,445 total TOPCAT patients, 1,767 were enrolled at American sites and 1,678 at Russia/Georgia sites; after geographic filtering and retaining patients with complete endpoint data, the American analysis included 719 patients (treatment and placebo arms combined).

The American-restricted (**Figure 5C** and **5D**) model preserved the core treatment-modifying variables while producing a substantially stronger heterogeneity signal. Diabetes mellitus (β_TX = −0.099), creatinine clearance below 30 mL/min (β_TX = +0.095), and bundle branch block (β_TX = +0.073) remained the top-ranked modifiers (Figure 5A, lower panel), supporting the cardiorenal phenotype as the principal axis of spironolactone response. Additional variables that gained prominence (e.g., edema, elevated jugular venous pressure, hypertension, and hematocrit) further reinforce a volume-overload signature as the key determinant of differential response. This volume overload variables is also a key finding pointing to the benefit of spironolactone for HFpEF patients with cardiorenal syndrome.

It is also worth noting that the treatment effect separation nearly doubled in the American cohort. Patients with predicted high ITE experienced a benefit of +5.8 KCCQ points, while those with predicted low ITE exhibited a treatment effect of −6.3 KCCQ points, yielding an interaction statistic of 12.1 (permutation P < 0.001). This result represents a 68% increase over the 7.2-point separation in the full cohort and is consistent with the hypothesis that the Russia/Georgia cohort introduced phenotypic noise, diluting the interaction signal. Additionally, halving the sample size was accompanied by a stronger P-value, which indicates the gain in signal purity more than compensated for the loss of statistical power.

#### NEAT-HFpEF (Isosorbide Mononitrate): Treatment-Modifying Variables

The NEAT-HFpEF ITE model identified a distinct hemodynamic phenotype. The top variables associated with treatment-modifying effect were pulse pressure (βTX = -0.022), rales (βTX = -0.021), systolic blood pressure (βTX = -0.017), insulin use (βTX = -0.016), and S3 gallop (βTX = -0.016), as shown in **Figure 6A**. The negative coefficients indicate that patients with higher pulse pressure, rales, and higher systolic blood pressure experienced less benefit (or greater harm) from isosorbide mononitrate. Mechanistically, these results are coherent since nitrate therapy primarily reduces preload^27^, and a patient who already has signs of congestion and high filling pressures may represent a phenotype in which preload reduction alone is insufficient or potentially harmful^41^. The baseline characteristics of patients in the treatment arm stratified by predicted ITE are described in **Table 5**.

**Figure 6.**
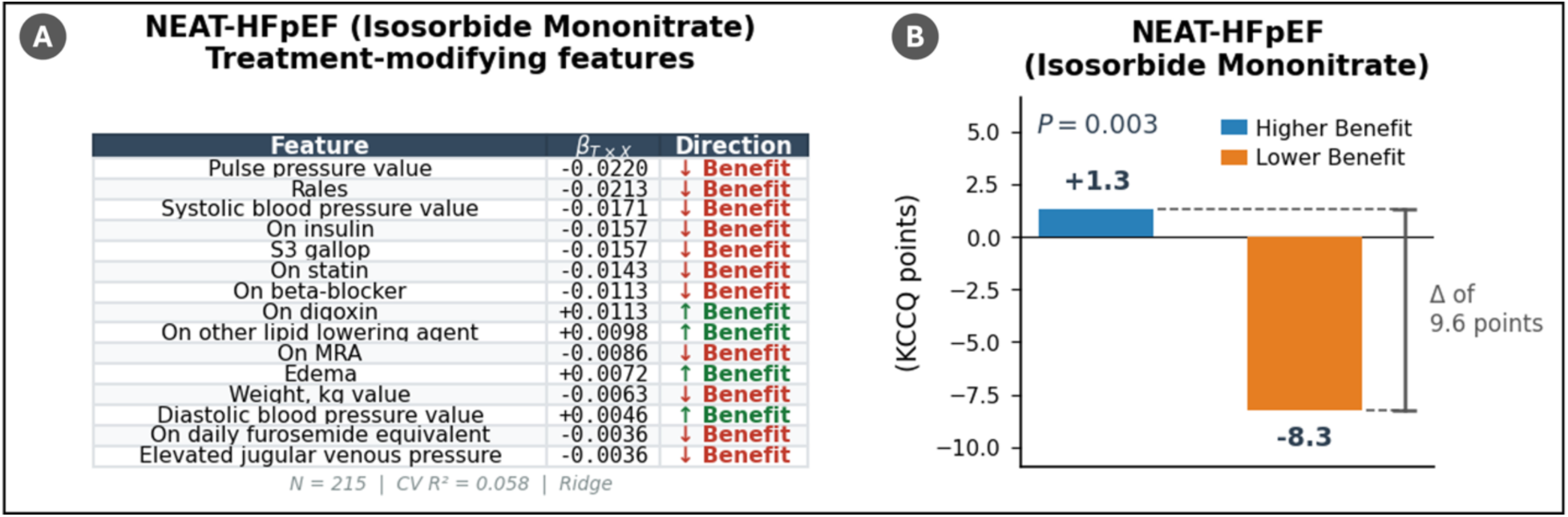
Interaction-based treatment effect model trained on NEAT-HFpEF (Isosorbide Mononitrate). (A) Non-zero treatment–covariate interaction coefficients (β_T×X) from the penalized Ridge regression model. Pulse pressure, rales, and systolic blood pressure emerged as the strongest interaction variables, all with negative coefficients suggesting that higher values are associated with reduced treatment benefit. An additional 6 variables are not shown. (B) Differential treatment effect across validation studies by predicted ITE subgroup. The NEAT-trained model identified a significant differential treatment effect when validated on its own cohort (NEAT: high ITE +1.3 vs. low ITE −8.3), with consistent directionality across external studies.

**Table 5.** Baseline characteristics of NEAT-HFpEF (Isosorbide Mononitrate) patients in the treatment arm stratified by predicted ITE median split. Patients classified as Higher Benefit (n = 54) versus Lower Benefit (n = 54) differed notably in hemodynamic profile and comorbidity burden. The Lower Benefit subgroup exhibited significantly higher systolic blood pressure (137.1 vs. 120.7 mmHg; SMD = 1.04) and pulse pressure (67.5 vs. 49.4 mmHg; SMD = 1.40), greater prevalence of diabetes mellitus (62.5% vs. 23.1%), higher BMI, and lower rates of beta-blocker (20.4% vs. 40.7%), statin (25.9% vs. 66.7%), and mineralocorticoid receptor antagonist use (64.8% vs. 83.3%). These differences suggest that patients with a wider pulse pressure and greater metabolic burden (i.e., variables reflecting arterial stiffness and systemic metabolic dysfunction) derive less benefit from nitrate therapy, consistent with the hypothesis that isosorbide mononitrate is more effective in patients with lower preload reserve rather than those with predominantly vascular or metabolic pathology.

| Characteristic | Lower Benefit (n=54) | Higher Benefit (n=54) | P Value | SMD |
| --- | --- | --- | --- | --- |
| **Demographics** |  |  |  |  |
| Age, years | 67.2 ± 8.0 | 71.7 ± 8.5 | **0.048** | 0.54 |
| Body mass index, kg/m<sup>2</sup> | 37.6 ± 9.0 | 34.1 ± 7.4 | **0.042** | 0.42 |
| Body surface area, m<sup>2</sup> | 2.2 ± 0.3 | 2.1 ± 0.3 | 0.218 | 0.45 |
| Female sex | 0 (0.0) | 0 (0.0) | 1.000 |  |
| Race |  |  | 0.848 |  |
| 1.0 | 20 (83.3) | 23 (88.5) |  |  |
| 2.0 | 3 (12.5) | 2 (7.7) |  |  |
| 6.0 | 1 (4.2) | 1 (3.8) |  |  |
| **Clinical Characteristics** |  |  |  |  |
| Systolic BP, mm Hg | 137.1 ± 16.9 | 120.7 ± 14.6 | **<0.001** | 1.04 |
| Diastolic BP, mm Hg | 69.6 ± 10.6 | 71.3 ± 11.7 | 0.394 | 0.15 |
| Pulse pressure, mm Hg | 67.5 ± 14.0 | 49.4 ± 11.7 | **<0.001** | 1.40 |
| HF duration, months | 1.3 (0.5–3.1) | 1.8 (0.9–4.7) | 0.336 | 0.33 |
| NYHA |  |  | 1.000 |  |
| 2.0 | 29 (53.7) | 28 (51.9) |  |  |
| 3.0 | 25 (46.3) | 26 (48.1) |  |  |
| **Comorbidities** |  |  |  |  |
| Diabetes mellitus | 15 (62.5) | 6 (23.1) | **0.011** | 0.80 |
| Atrial fibrillation | 7 (100.0) | 12 (100.0) | 1.000 |  |
| Prior MI | 2 (8.3) | 4 (15.4) | 0.669 | 0.22 |
| Prior CABG | 0 (0.0) | 5 (19.2) | 0.051 | 0.64 |
| COPD | 3 (12.5) | 3 (11.5) | 1.000 | 0.03 |
| CKD | 11 (45.8) | 11 (44.0) | 1.000 | 0.04 |
| Prior stroke | 2 (8.3) | 1 (3.8) | 0.602 | 0.19 |
| PVD | 2 (8.3) | 1 (4.0) | 0.609 | 0.18 |
| **Laboratory Values** |  |  |  |  |
| Hemoglobin, g/dL | 13.1 ± 1.4 | 13.0 ± 1.3 | 0.928 | 0.13 |
| Hematocrit, % | 39.9 ± 3.5 | 39.3 ± 3.9 | 0.741 | 0.17 |
| Potassium, mEq/L | 4.2 ± 0.6 | 4.4 ± 0.4 | 0.102 | 0.36 |
| BUN, mg/dL | 28.9 ± 20.3 | 20.3 ± 5.1 | 0.609 | 0.59 |
| CrCl, mL/min | 61.6 ± 24.2 | 64.0 ± 20.1 | 0.575 | 0.11 |
| Glucose, mg/dL | 117.9 ± 46.9 | 107.5 ± 37.0 | 0.264 | 0.25 |
| NT-proBNP/BNP (harmonized) | 258.4 (88.7–691.5) | 178.2 (108.1–515.6) | 0.564 | 0.06 |
| **Echocardiography** |  |  |  |  |
| LVEF, % | 61.0 ± 6.3 | 59.1 ± 9.2 | 0.631 | 0.24 |
| LV mass index, g/m<sup>2</sup> | 82.5 ± 27.4 | 81.3 ± 21.1 | 0.957 | 0.05 |
| LA volume index, mL/m<sup>2</sup> | 44.2 ± 21.9 | 40.9 ± 17.4 | 0.635 | 0.17 |
| E/e' ratio | 12.8 ± 5.5 | 14.3 ± 10.7 | 0.894 | 0.18 |
| RVSP, mm Hg | 34.7 ± 17.4 | 30.5 ± 6.8 | 0.901 | 0.33 |
| **Quality of Life** |  |  |  |  |
| KCCQ Overall Summary | 58.8 ± 25.3 | 59.3 ± 23.9 | 0.968 | 0.02 |
| KCCQ Clinical Summary | 58.8 ± 24.1 | 61.1 ± 21.8 | 0.696 | 0.10 |
| KCCQ Physical Limitation | 56.7 ± 25.7 | 61.3 ± 24.4 | 0.317 | 0.18 |
| **Medications** |  |  |  |  |
| ACEI/ARB | 0 (0.0) | 0 (0.0) | 1.000 |  |
| Beta-blocker | 11 (20.4) | 22 (40.7) | **0.037** | 0.44 |
| CCB | 35 (64.8) | 39 (72.2) | 0.534 | 0.16 |
| MRA | 35 (64.8) | 45 (83.3) | **0.048** | 0.42 |
| Loop diuretic | 0 (0.0) | 0 (0.0) | 1.000 |  |
| Statin | 14 (25.9) | 36 (66.7) | **<0.001** | 0.82 |
| Aspirin | 0 (0.0) | 0 (0.0) | 1.000 |  |
| Anticoagulant | 0 (0.0) | 0 (0.0) | 1.000 |  |

NEAT-HFpEF (Isosorbide Mononitrate) — Treatment arm, ITE median split. Values are mean ± SD, median (IQR), or n (%). P: Mann-Whitney U or Chi-square/Fisher. SMD = standardized mean difference.

Although the NEAT-HFpEF trial reported that isosorbide mononitrate reduced daily activity levels compared with placebo in patients with HFpEF, leading to the conclusion that nitrates are ineffective or harmful in this population, the ITE model revealed significant heterogeneity in treatment response. Our validation showed a significant interaction of +9.6 (permutation p=0.003), suggesting that the model distinguished patients based on treatment benefit **(Figure 6B)**. The predicted high-ITE subgroup improved with isosorbide mononitrate (treatment: +5.4 vs. placebo: +4.1; TE=+1.3). This group is characterized by patients with lower congestion burden (lower pulse pressure, fewer rales, lower systolic BP, no elevated JVP) and less intensive background medical therapy (not on insulin, beta-blockers, statins, or MRAs). This suggests that less hemodynamically compromised HFpEF patients derive greater benefit from isosorbide mononitrate. In contrast, the low-ITE subgroup declined with treatment (treatment: −4.2 vs. placebo: +4.1; TE=−8.3) and comprises patients with higher congestion burden (elevated pulse pressure, rales, elevated JVP) and more intensive background therapy (on insulin, beta-blockers, statins, MRAs, higher furosemide doses). These are more hemodynamically compromised patients in whom isosorbide mononitrate appears to worsen quality of life, likely through hypotension and reduced activity tolerance. In other words, the high- and low-ITE groups represent, respectively, subgroups that showed modest benefit and substantial decline with treatment respectively, similarly to the TOPCAT results. When analyzed all together, these opposing effects could explain the overall net negative signal reported in the original trial analysis. Although further validation in external cohorts and prospective analyses is needed, if confirmed, these findings have implications for the design and interpretation of HFpEF trials, where phenotypic heterogeneity should be considered, as it may mask meaningful treatment effects in certain patient subgroups.

#### INDIE-HFpEF (Inorganic Nitrite): Treatment-Modifying Variables

The ITE model trained on the INDIE-HFpEF trial identified variables associated with volume overload and metabolic function as top treatment modifiers. These variables were BUN (βTX = -0.033), elevated jugular venous pressure (βTX = -0.028), daily furosemide equivalent dose (βTX = -0.024), diastolic blood pressure (βTX = -0.023), and glucose (βTX = -0.021) as illustrated in **Figure 7A**. This profile suggests that patients with more advanced volume overload and metabolic derangement derive less benefit from inorganic nitrite supplementation. This is consistent with evidence from previous work showing that comorbidities such as obesity, diabetes, and chronic kidney disease impair the NO-cGMP-PKG signaling axis at multiple levels through oxidative stress and endothelial inflammation^42^. This impairment creates a state in which exogenous NO supplementation alone cannot restore downstream pathway function^26^. Furthermore, in patients with established volume overload, the steep diastolic pressure-volume relationship characteristic of HFpEF limits the hemodynamic benefit of preload reduction^28^. Internal validation (**Figure 7B**) using cross-validated ITE predictions yielded a significant interaction of +11.2 (permutation p=0.004), with predicted high-ITE patients showing a treatment effect of +4.9 (treatment: +5.9, placebo: +1.0) compared to −6.3 in low-ITE patients (treatment: −1.5, placebo: +4.8). The high-ITE group includes leaner patients with better renal function (lower BUN, higher creatinine clearance, lower CKD stage) and less congestion (no elevated JVP, lower furosemide doses), while the low-ITE group is characterized by greater cardiorenal and metabolic burden, suggesting that impaired renal function and volume overload may limit nitrite-to-NO bioconversion and blunt therapeutic benefit. The baseline characteristics of patients in the treatment arm stratified by predicted ITE are described in **Table 6**.

**Figure 7.**
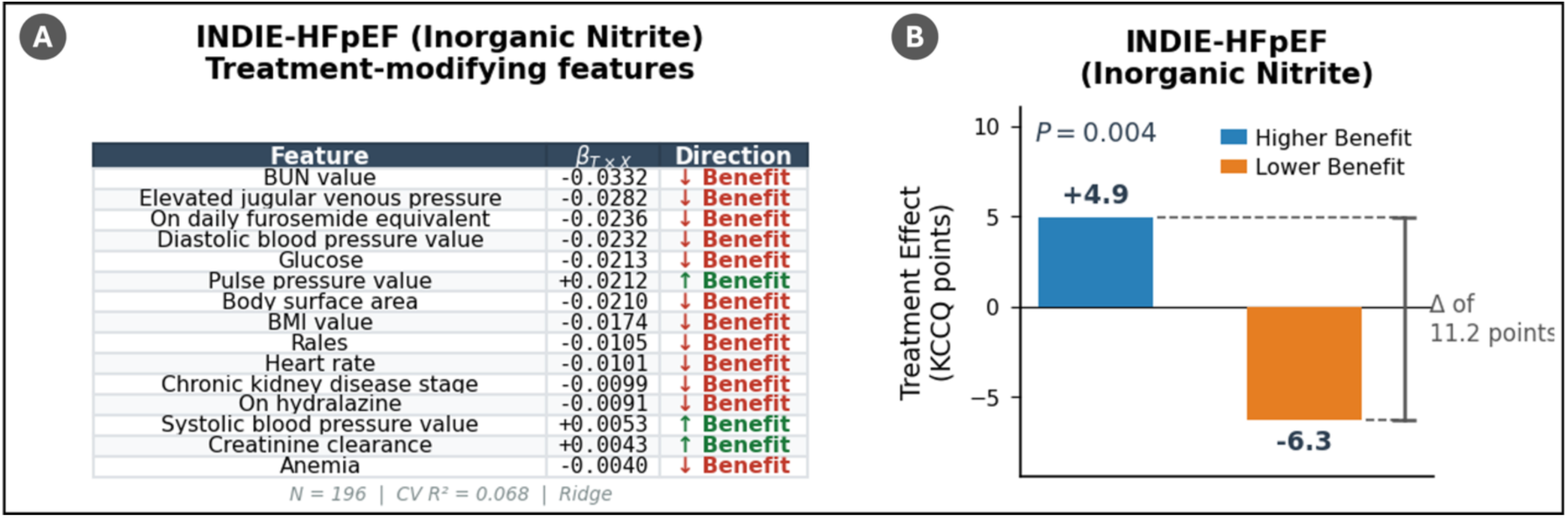
Interaction-based treatment effect model trained on INDIE-HFpEF (Inorganic Nitrite). (A) Non-zero treatment–covariate interaction coefficients (β_T×X). BUN value was the dominant interaction variable, with elevated BUN associated with reduced predicted treatment benefit. Pulse pressure value was the only variable with a positive interaction coefficient of notable magnitude. One additional variable is not shown. (B) Differential treatment effect across validation studies by predicted ITE subgroup. The INDIE-trained model identified patients with a predicted treatment benefit of +4.9 in its own cohort, compared to −6.3 in the predicted low-ITE subgroup.

**Table 6.** Baseline characteristics of INDIE-HFpEF (Inorganic Nitrite) patients in the treatment arm stratified by predicted ITE median split. Patients classified as Higher Benefit (n = 50) versus Lower Benefit (n = 49) differed significantly in age (71.0 vs. 64.8 years; SMD = 0.77), body habitus (BMI 32.5 vs. 37.3 kg/m²; SMD = 0.81; BSA 2.0 vs. 2.2 m²; SMD = 0.94), and renal function, with the Lower Benefit subgroup exhibiting markedly higher BUN (29.0 vs. 18.5 mg/dL; SMD = 0.91), lower creatinine clearance (54.7 vs. 66.2 mL/min; SMD = 0.61), and higher resting heart rate. The Lower Benefit group also had lower rates of beta-blocker use (22.4% vs. 46.0%) and a trend toward greater diabetes prevalence (45.8% vs. 29.6%) and higher LV mass index..

| Characteristic | Lower Benefit (n=49) | Higher Benefit (n=50) | P Value | SMD |
| --- | --- | --- | --- | --- |
| **Demographics** |  |  |  |  |
| Age, years | 64.8 ± 9.2 | 71.0 ± 6.7 | **0.010** | 0.77 |
| Body mass index, kg/m<sup>2</sup> | 37.3 ± 5.4 | 32.5 ± 6.4 | **<0.001** | 0.81 |
| Body surface area, m<sup>2</sup> | 2.2 ± 0.2 | 2.0 ± 0.2 | **<0.001** | 0.94 |
| Female sex | 0 (0.0) | 0 (0.0) | 1.000 |  |
| Race |  |  | 0.890 |  |
| 1.0 | 21 (87.5) | 25 (92.6) |  |  |
| 2.0 | 3 (12.5) | 2 (7.4) |  |  |
| **Clinical Characteristics** |  |  |  |  |
| Systolic BP, mm Hg | 127.4 ± 16.6 | 131.2 ± 17.4 | 0.186 | 0.22 |
| Diastolic BP, mm Hg | 74.3 ± 11.3 | 71.1 ± 10.2 | 0.080 | 0.30 |
| Pulse pressure, mm Hg | 53.1 ± 15.6 | 60.1 ± 16.1 | **0.010** | 0.44 |
| HF duration, months | 30.7 (10.2-55.7) | 16.6 (4.0-56.5) | 0.218 | 0.02 |
| **Comorbidities** |  |  |  |  |
| Diabetes mellitus | 11 (45.8) | 8 (29.6) | 0.366 | 0.34 |
| Prior MI | 0 (0.0) | 1 (3.7) | 1.000 | 0.27 |
| Prior CABG | 2 (8.3) | 2 (7.4) | 1.000 | 0.03 |
| COPD | 1 (4.2) | 3 (11.1) | 0.612 | 0.26 |
| CKD | 17 (34.7) | 27 (55.1) | 0.068 | 0.41 |
| Prior stroke | 1 (4.2) | 2 (7.4) | 1.000 | 0.14 |
| PVD | 1 (4.2) | 2 (7.4) | 1.000 | 0.14 |
| **Laboratory Values** |  |  |  |  |
| Hemoglobin, g/dL | 13.0 ± 1.7 | 13.1 ± 1.4 | 0.795 | 0.05 |
| Hematocrit, % | 39.2 ± 4.2 | 39.8 ± 3.5 | 0.430 | 0.17 |
| Sodium, mEq/L | 139.8 ± 2.9 | 140.6 ± 2.2 | 0.213 | 0.30 |
| BUN, mg/dL | 29.0 ± 14.9 | 18.5 ± 6.6 | **<0.001** | 0.91 |
| CrCl, mL/min | 54.7 ± 17.5 | 66.2 ± 20.0 | **0.002** | 0.61 |
| Glucose, mg/dL | 129.6 ± 59.8 | 112.1 ± 40.1 | 0.051 | 0.34 |
| **Echocardiography** |  |  |  |  |
| LVEF, % | 60.9 ± 5.5 | 62.1 ± 4.6 | 0.409 | 0.24 |
| LV mass index, g/m<sup>2</sup> | 107.0 ± 28.4 | 92.3 ± 28.4 | 0.081 | 0.52 |
| LA volume index, mL/m<sup>2</sup> | 42.5 ± 19.1 | 40.1 ± 24.0 | 0.384 | 0.11 |
| E/e' ratio | 15.1 ± 6.8 | 16.1 ± 10.4 | 0.990 | 0.11 |
| RVSP, mm Hg | 41.2 ± 13.5 | 38.3 ± 14.0 | 0.595 | 0.22 |
| **Quality of Life** |  |  |  |  |
| KCCQ Overall Summary | 57.0 ± 17.2 | 62.0 ± 18.2 | 0.206 | 0.28 |
| KCCQ Clinical Summary | 57.9 ± 17.1 | 64.6 ± 17.8 | 0.093 | 0.39 |
| KCCQ Physical Limitation | 58.1 ± 18.7 | 63.5 ± 22.6 | 0.138 | 0.26 |
| **Medications** |  |  |  |  |
| ACEi/ARB | 0 (0.0) | 0 (0.0) | 1.000 |  |
| Beta-blocker | 11 (22.4) | 23 (46.0) | **0.024** | 0.50 |
| CCB | 38 (77.6) | 34 (68.0) | 0.400 | 0.21 |
| MRA | 30 (61.2) | 37 (74.0) | 0.253 | 0.27 |
| Loop diuretic | 0 (0.0) | 0 (0.0) | 1.000 |  |
| Aspirin | 0 (0.0) | 0 (0.0) | 1.000 |  |
| Anticoagulant | 27 (55.1) | 33 (66.0) | 0.366 | 0.22 |

INDIE-HFpEF (Inorganic Nitrite) — Treatment arm, ITE median split. Values are mean ± SD, median (IQR), or n (%). P: Mann-Whitney U or Chi-square/Fisher. SMD = standardized mean difference.

#### RELAX (Sildenafil): Treatment-Modifying Variables

The RELAX ITE model identified a different interaction profile associated with inflammation and volume overload markers. As shown in **Figure 8A**, chronic kidney disease stage, C-reactive protein, edema, creatinine clearance <30, and aortic regurgitation are the top treatment-modifying variables for this model. Despite a treatment effect differential of +0.8 VO2 points between subgroups (**Figure 8B**), the interaction did not reach statistical significance (permutation p=0.10), likely reflecting both the smaller sample size (n=189) and the use of peak VO2 as the endpoint, which operates on a narrower scale than KCCQ. Although treatment effect heterogeneity for sildenafil in HFpEF may exist, the current data are insufficient to reliably characterize treatment-modifying variables. The baseline characteristics of patients in the treatment arm stratified by predicted ITE are described in **Table 7**.

**Figure 8.**
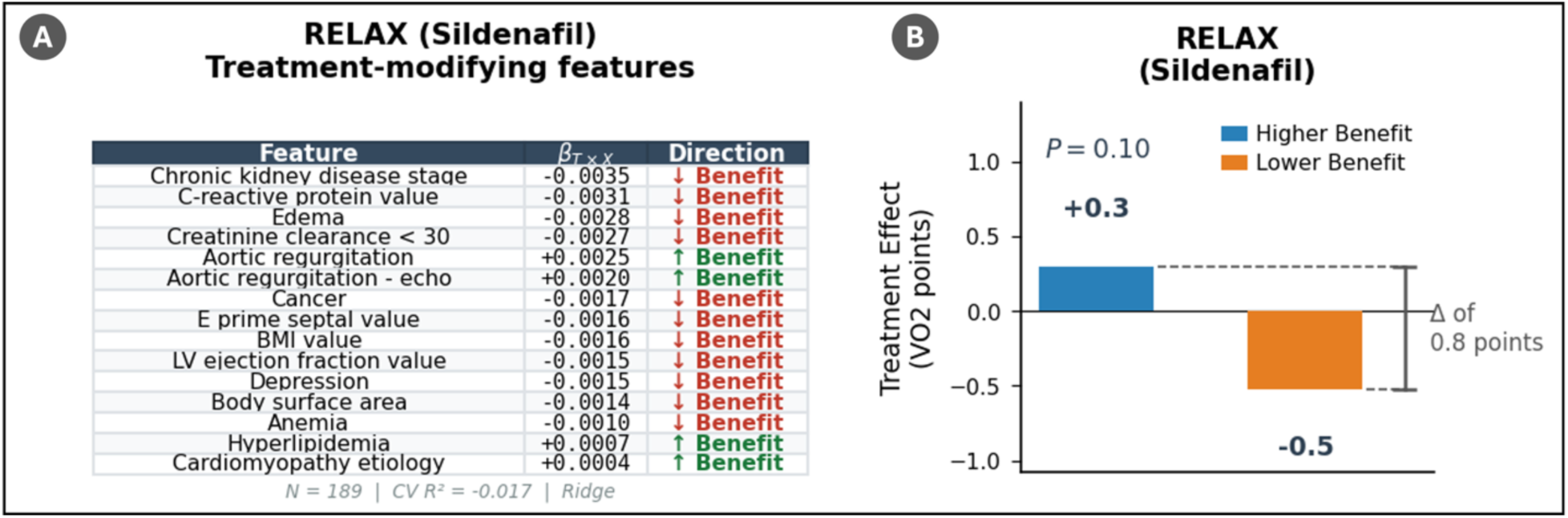
Interaction-based treatment effect model trained on RELAX (Sildenafil). (A) Non-zero treatment–covariate interaction coefficients (β_T×X). Chronic kidney disease stage was the strongest negative interaction variable, while aortic regurgitation showed the largest positive interaction, suggesting that patients with aortic regurgitation may derive relatively greater benefit from sildenafil. An additional variables not shown. (B) Differential treatment effect by predicted ITE subgroup. The RELAX model showed a +0.8 VO2-point differential between high-ITE (+0.3) and low-ITE (−0.5) subgroups, but this did not reach statistical significance (p=0.10), likely due to the small sample size (N=189) and the narrow dynamic range of peak VO2.

**Table 7.** Baseline characteristics of RELAX (Sildenafil) patients in the treatment arm stratified by predicted ITE median split. Patients classified as Higher Benefit (n = 54) versus Lower Benefit (n = 53) differed most prominently in body habitus, with the Lower Benefit subgroup exhibiting significantly higher BMI (37.3 vs. 31.5 kg/m²; SMD = 0.86) and body surface area (2.2 vs. 2.0 m²; SMD = 1.18). The Lower Benefit group also showed higher resting heart rate, more advanced NYHA class (62.3% vs. 40.7% NYHA III), lower creatinine clearance (53.8 vs. 62.4 mL/min), and higher E/e′ ratio (14.4 vs. 18.0). This result suggest an overlap with the obesity-related, hemodynamically congested phenotype associated with attenuated response.

| Characteristic | Lower Benefit (n=53) | Higher Benefit (n=54) | P Value | SMD |
| --- | --- | --- | --- | --- |
| **Demographics** |  |  |  |  |
| Age, years | 67.6 ± 10.7 | 68.9 ± 10.1 | 0.449 | 0.13 |
| Body mass index, kg/m² | 37.3 ± 7.5 | 31.5 ± 5.8 | <0.001 | 0.86 |
| Body surface area, m² | 2.2 ± 0.2 | 2.0 ± 0.2 | <0.001 | 1.18 |
| Female sex | 0 (0.0) | 0 (0.0) | 1.000 |  |
| Race |  |  | 0.097 |  |
| 1.0 | 49 (92.5) | 48 (88.9) |  |  |
| 2.0 | 0 (0.0) | 4 (7.4) |  |  |
| 6.0 | 4 (7.5) | 2 (3.7) |  |  |
| **Clinical Characteristics** |  |  |  |  |
| Systolic BP, mm Hg | 123.4 ± 15.8 | 131.2 ± 20.0 | 0.070 | 0.43 |
| Diastolic BP, mm Hg | 70.3 ± 9.7 | 70.7 ± 10.7 | 0.779 | 0.04 |
| Pulse pressure, mm Hg | 53.1 ± 13.9 | 60.4 ± 19.4 | 0.073 | 0.44 |
| HF duration, months | 24.7 (3.7–58.1) | 18.8 (4.9–41.7) | 0.769 | 0.12 |
| NYHA |  |  | 0.042 |  |
| 2.0 | 20 (37.7) | 32 (59.3) |  |  |
| 3.0 | 33 (62.3) | 22 (40.7) |  |  |
| **Comorbidities** |  |  |  |  |
| Diabetes mellitus | 28 (52.8) | 18 (33.3) | 0.066 | 0.39 |
| Atrial fibrillation | 32 (60.4) | 24 (44.4) | 0.145 | 0.32 |
| Prior MI | 9 (100.0) | 6 (100.0) | 1.000 |  |
| Prior CABG | 7 (100.0) | 9 (100.0) | 1.000 |  |
| COPD | 12 (22.6) | 7 (13.0) | 0.291 | 0.25 |
| CKD | 17 (32.1) | 22 (40.7) | 0.465 | 0.18 |
| Prior stroke | 3 (5.7) | 4 (7.4) | 1.000 | 0.07 |
| PVD | 9 (17.0) | 7 (13.0) | 0.755 | 0.11 |
| **Laboratory Values** |  |  |  |  |
| Hemoglobin, g/dL | 12.8 ± 1.6 | 13.2 ± 1.5 | 0.237 | 0.26 |
| Hematocrit, % | 37.7 ± 4.6 | 38.9 ± 4.3 | 0.237 | 0.26 |
| Sodium, mEq/L | 140.1 ± 2.8 | 139.7 ± 3.4 | 0.389 | 0.13 |
| Potassium, mEq/L | 4.3 ± 0.6 | 4.4 ± 0.5 | 0.574 | 0.15 |
| BUN, mg/dL | 29.3 ± 14.6 | 25.7 ± 11.3 | 0.270 | 0.27 |
| CrCl, mL/min | 53.8 ± 20.2 | 62.4 ± 19.5 | 0.032 | 0.43 |
| NT-proBNP/BNP (harmonized) | 1420.0 (693.0–2389.5) | 1170.0 (439.0–2051.0) | 0.216 | 0.23 |
| **Echocardiography** |  |  |  |  |
| LVEF, % | 61.0 ± 6.5 | 61.1 ± 6.8 | 0.960 | 0.03 |
| LA volume index, mL/m² | 49.6 ± 21.2 | 52.2 ± 29.4 | 0.995 | 0.10 |
| E/e' ratio | 14.4 ± 7.9 | 18.0 ± 11.3 | 0.043 | 0.36 |
| RVSP, mm Hg | 40.7 ± 11.7 | 44.1 ± 12.0 | 0.377 | 0.28 |
| **Quality of Life** |  |  |  |  |
| 6MWD, m | 291.9 ± 131.9 | 337.6 ± 87.3 | 0.092 | 0.41 |
| **Medications** |  |  |  |  |
| Loop diuretic | 0 (0.0) | 0 (0.0) | 1.000 |  |

RELAX (Sildenafil) — Treatment arm, ITE median split. Values are mean ± SD, median (IQR), or n (%). P: Mann-Whitney U or Chi-square/Fisher. SMD = standardized mean difference.

#### Cross-Trial Validation: Evidence for Drug Specificity

The cross-trial validation matrix (**Figure 9**) provides evidence that supports the validity of the proposed ITE approach. When each trial’s ITE model was applied to all four trials, a consistent pattern of diagonal significance emerged: each model produced a statistically significant interaction only when validated on its own trial’s drug. The TOPCAT model was significant only for TOPCAT (p=0.001; RELAX p=0.265, NEAT-HFpEF p=0.419, INDIE-HFpEF p=0.622). The NEAT-HFpEF model was significant only for NEAT-HFpEF (p=0.003; TOPCAT p=0.099, RELAX p=0.409, INDIE-HFpEF p=0.778). The lack of interactions between organic nitrate (NEAT-HF) and inorganic nitrite (INDIE-HFpEF) may be expected due to significant differences in the mechanisms of action, with regard to differences in bioavailability and development of tolerance and possibly reflex neurohormonal activation with nitrates. In contrast, inorganic nitrite provides NO during exercise in muscles where oxygen tension are low. The INDIE-HFpEF model was significant for INDIE-HFpEF (p=0.004) and also showed a significant interaction when validated on RELAX (p=0.036), which may reflect shared vasodilatory mechanisms between inorganic nitrite and sildenafil, as both act through nitric oxide signaling pathways. The RELAX model did not reach significance on any trial (self-validation p=0.104), though the TOPCAT validation approached significance (p=0.059), consistent with the limited statistical power discussed above. The one notable off-diagonal signal, between INDIE-HFpEF and RELAX, is consistent with their shared distal action on the oxide–cGMP axis^43,44^, and their shared property to reduce oxidative stress.^45,46^ NEAT-HFpEF acts more upstream with distinct pharmacology,^47^ but paradoxically increases oxidative stress and induces endothelial dysfunction, which may explain its absence.^48,49^ This should be interpreted cautiously, as the RELAX model did not pass the permutation test, its cross-trial transportability is itself uncertain, and the apparent mechanistic coherence could reflect chance.

**Figure 9.**
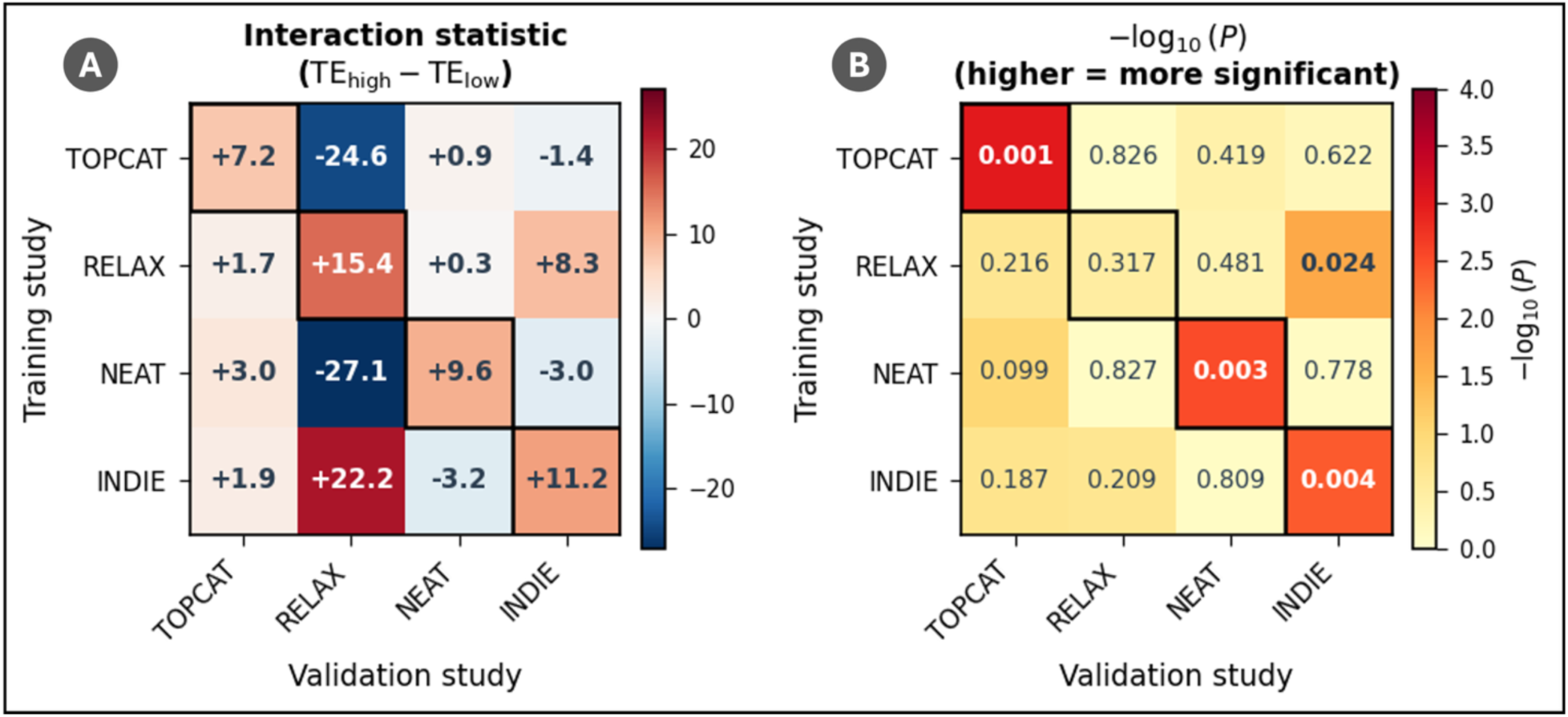
Cross-validation matrix of interaction-based treatment effect models across HFpEF clinical trials. (A) Interaction statistic (TE_high − TE_low) for each combination of training study (rows) and validation study (columns). Positive values indicate that the predicted high-ITE subgroup experienced a greater treatment effect than the predicted low-ITE subgroup. Diagonal cells with bold borders represent internal validation. The largest interaction statistics consistently appear along the diagonal, indicating that each model’s treatment-modifying signal is strongest for its own drug. (B) Permutation test p-values displayed as −log₁₀(P) for each training–validation pair. Statistically significant interactions (P < 0.05, shown in bold red) were identified for TOPCAT validated on itself (P = 0.001), NEAT-HFpEF validated on itself (P = 0.003), INDIE-HFpEF validated on itself (P = 0.004), and the INDIE-trained model validated on RELAX (P = 0.036). This diagonal-dominant pattern supports the interpretation that the ITE models capture drug-specific rather than generic prognostic signals.

This diagonal pattern provides compelling evidence that the ITE models capture drug-specific treatment effect modification rather than a generic “treatment responsiveness” phenotype. If the models were identifying a general propensity to improve, as observed in the prognostic model presented earlier, they would transfer across trials. Instead, the cardiorenal variables that modify spironolactone response do not predict differential response to nitrates, and the hemodynamic variables that modify nitrate response do not predict differential response to spironolactone.

ITE-outcome correlations within treatment arms further supported this specificity. In each trial, the ITE scores from the trial’s own model correlated significantly with observed outcomes in the treatment arm (TOPCAT: ρ=0.205, p<0.001; RELAX: ρ=0.221, p=0.032; NEAT-HFpEF: ρ=0.380, p<0.001; INDIE-HFpEF: ρ=0.393, p<0.001), whereas cross-trial ITE scores showed no significant correlation.

#### Surrogate Decision Trees for Clinical Interpretability

To translate the continuous ITE predictions into clinically actionable decision rules, surrogate decision trees (depth=3) were trained to approximate each trial’s ITE model. As mentioned before, these surrogate trees approximate the ITE model behavior, and a high level of fidelity is vital to ensure their results match the ITE. In this context, the TOPCAT tree achieved relatively high fidelity to the ITE model with an R² of 0.665 (ρ=0.771, agreement=78.8%). Using this tree, we identified diabetes mellitus as the root split, followed by bundle branch block and oral hypoglycemic use (**Figure 10**). Similar results were observed in the NEAT-HFpEF tree, which achieved an R² of 0.642 (ρ=0.815, agreement=82.3%). In this case, insulin use was used as the root of the tree, with pulse pressure and statin use as secondary splits (**Figure 11**). Next, the INDIE-HFpEF tree reached an R² of 0.587 (ρ=0.724, agreement=77.6%) and identified BUN as the root split, followed by body surface area and BMI (**Figure 12**). Lastly, the surrogate tree for the RELAX ITE model reached an R² of 0.540 (ρ=0.742, agreement=80.6%). The RELAX surrogate tree used edema as the root split, followed by body surface area and LV ejection fraction (**Figure 13**), reflecting a volume-overload and hemodynamic phenotype consistent with sildenafil’s mechanism of action. Together, these decision trees provide a more interpretable, bedside-applicable set of rules for phenotype-guided treatment selection.

**Figure 10.**
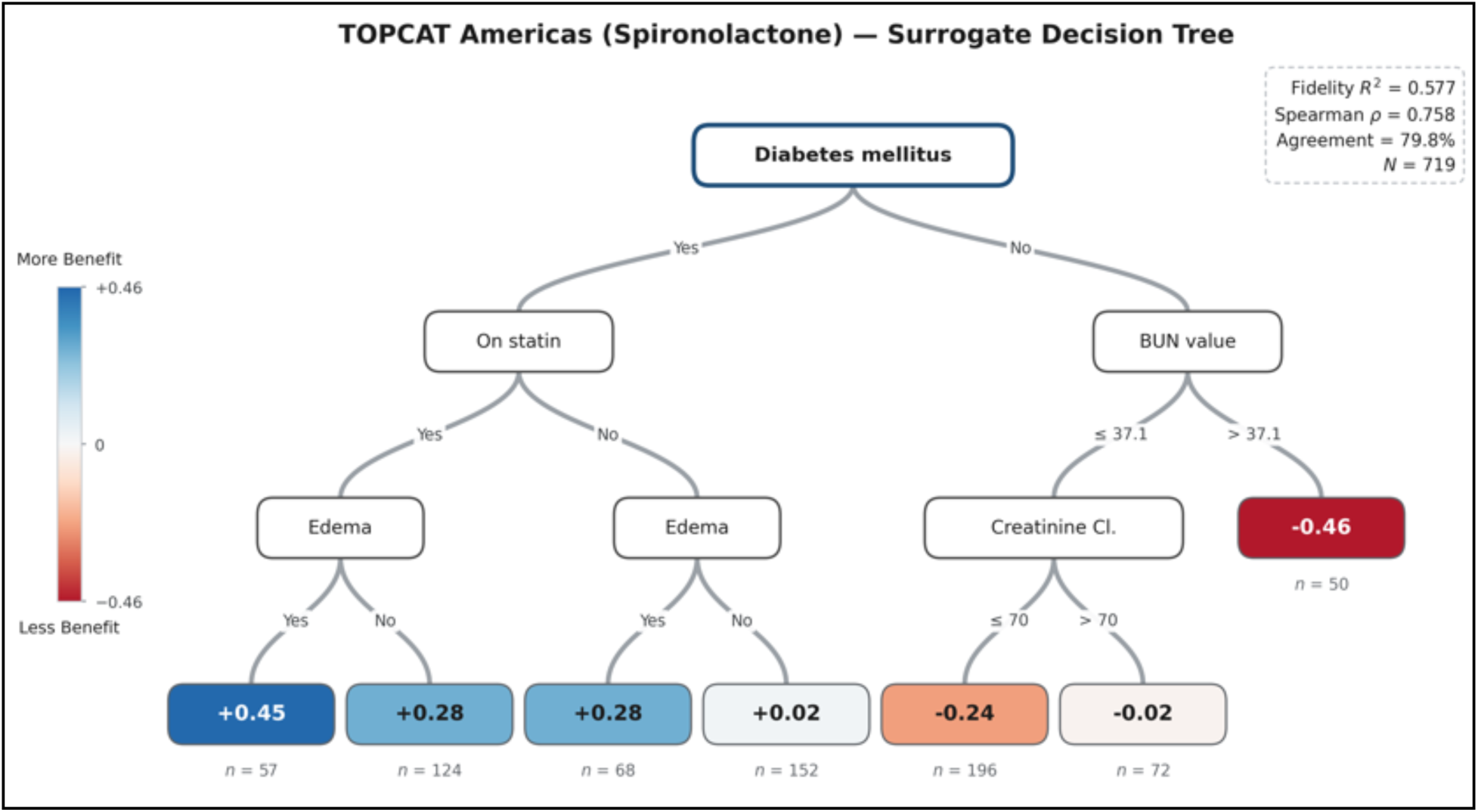
Surrogate decision tree approximating the TOPCAT-trained interaction model for spironolactone treatment effect prediction. A depth-3 decision tree was fit to approximate the complex Ridge interaction model’s individualized treatment effect (ITE) predictions using un-standardized baseline variables. Diabetes mellitus was the primary split: patients without diabetes were predicted to benefit from spironolactone (ITE range: +0.06 to +0.50), with the greatest benefit among those with bundle branch block (Any BBB) who were not on statins (+0.50). Patients with diabetes were predicted to experience harm, particularly those on oral hypoglycemics with low creatinine clearance (≤71.9 mL/min; ITE = −0.57). Leaf node colors indicate predicted treatment effect direction and magnitude (blue = benefit, red = harm). Surrogate fidelity: R² = 0.665, Spearman ρ = 0.771, high/low agreement = 78.8%.

**Figure 11.**
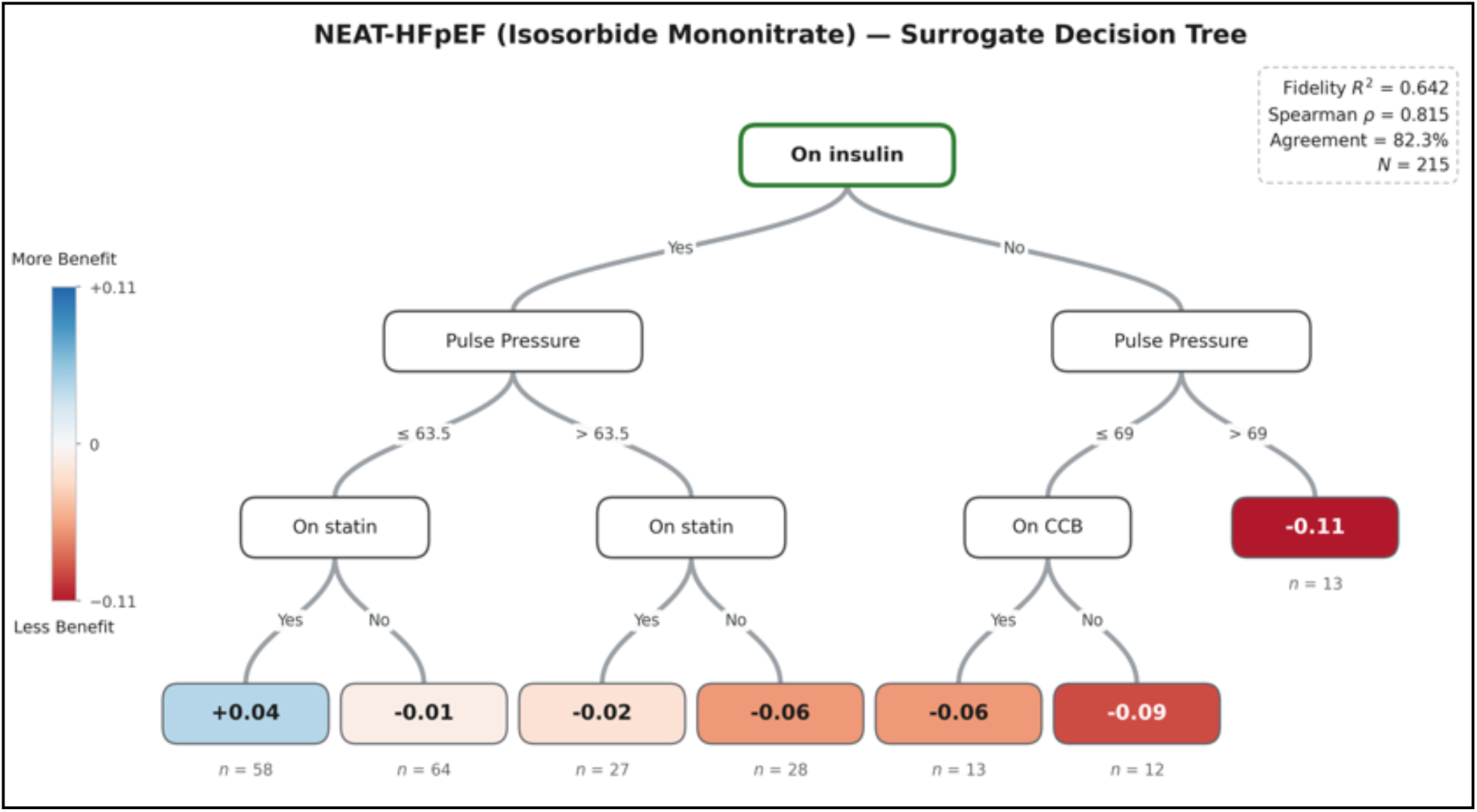
Surrogate decision tree approximating the NEAT-HFpEF–trained interaction model for isosorbide mononitrate treatment effect prediction. Insulin use was the primary split, with insulin-dependent patients uniformly predicted to experience harm (ITE range: −0.06 to −0.11). Among non-insulin patients, pulse pressure further stratified outcomes: those with lower pulse pressure (≤63.5 mmHg) who were not on statins showed the only predicted benefit (+0.04). The narrow ITE range (−0.11 to +0.04) reflects the overall modest heterogeneity of treatment effect in this trial. Surrogate fidelity: R² = 0.642, Spearman ρ = 0.815, high/low agreement = 82.3%.

**Figure 12.**
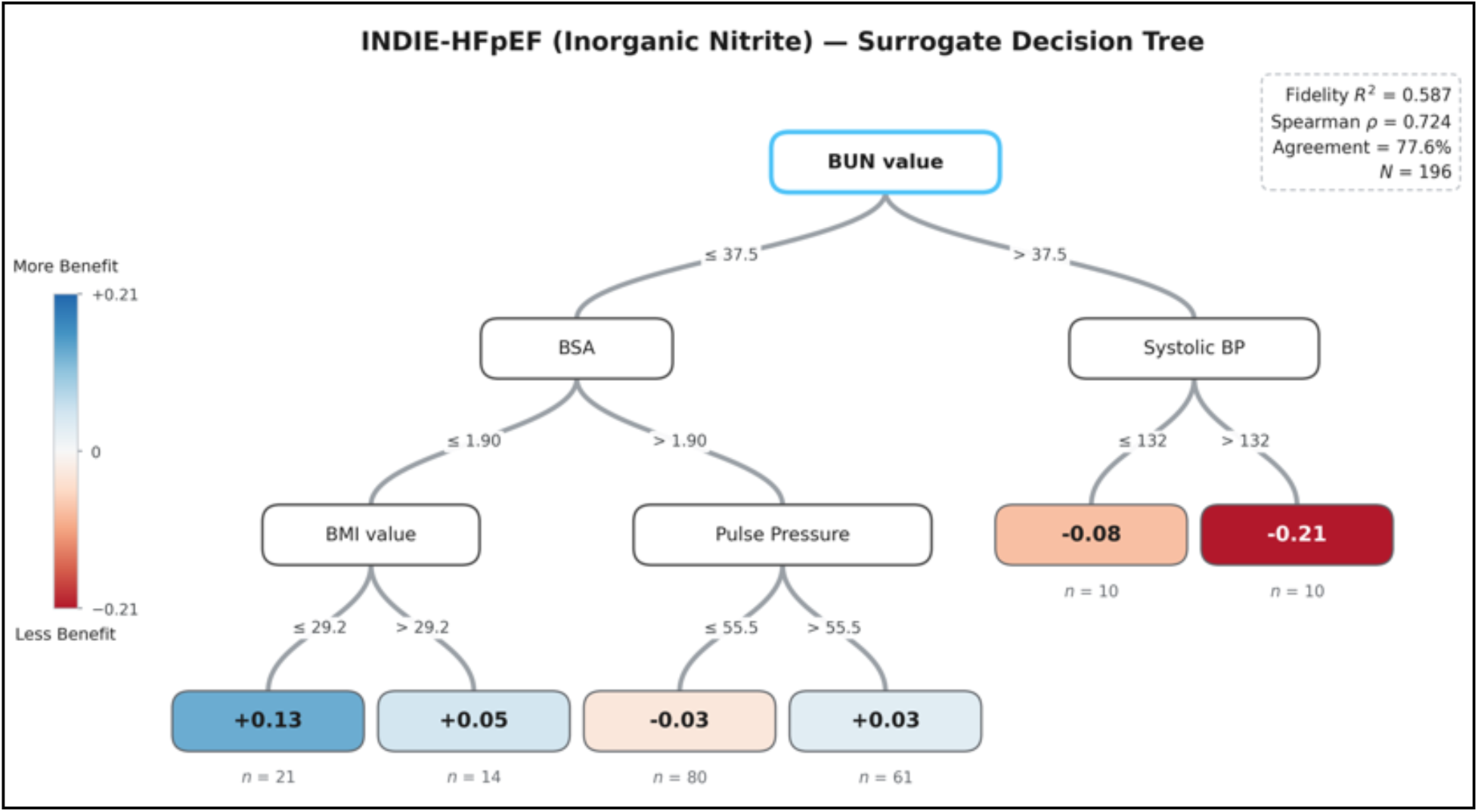
Surrogate decision tree approximating the INDIE-HFpEF–trained interaction model for inorganic nitrite treatment effect prediction. BUN value was the primary split: patients with elevated BUN (>37.5 mg/dL) were uniformly predicted to experience harm, with higher systolic blood pressure (>132 mmHg) further worsening the predicted effect (ITE = −0.21). Among patients with lower BUN, smaller body surface area (≤1.90 m²) and lower BMI (≤29.24 kg/m²) were associated with the greatest predicted benefit (ITE = +0.13). Surrogate fidelity: R² = 0.587, Spearman ρ = 0.724, high/low agreement = 77.6%.

**Figure 13.**
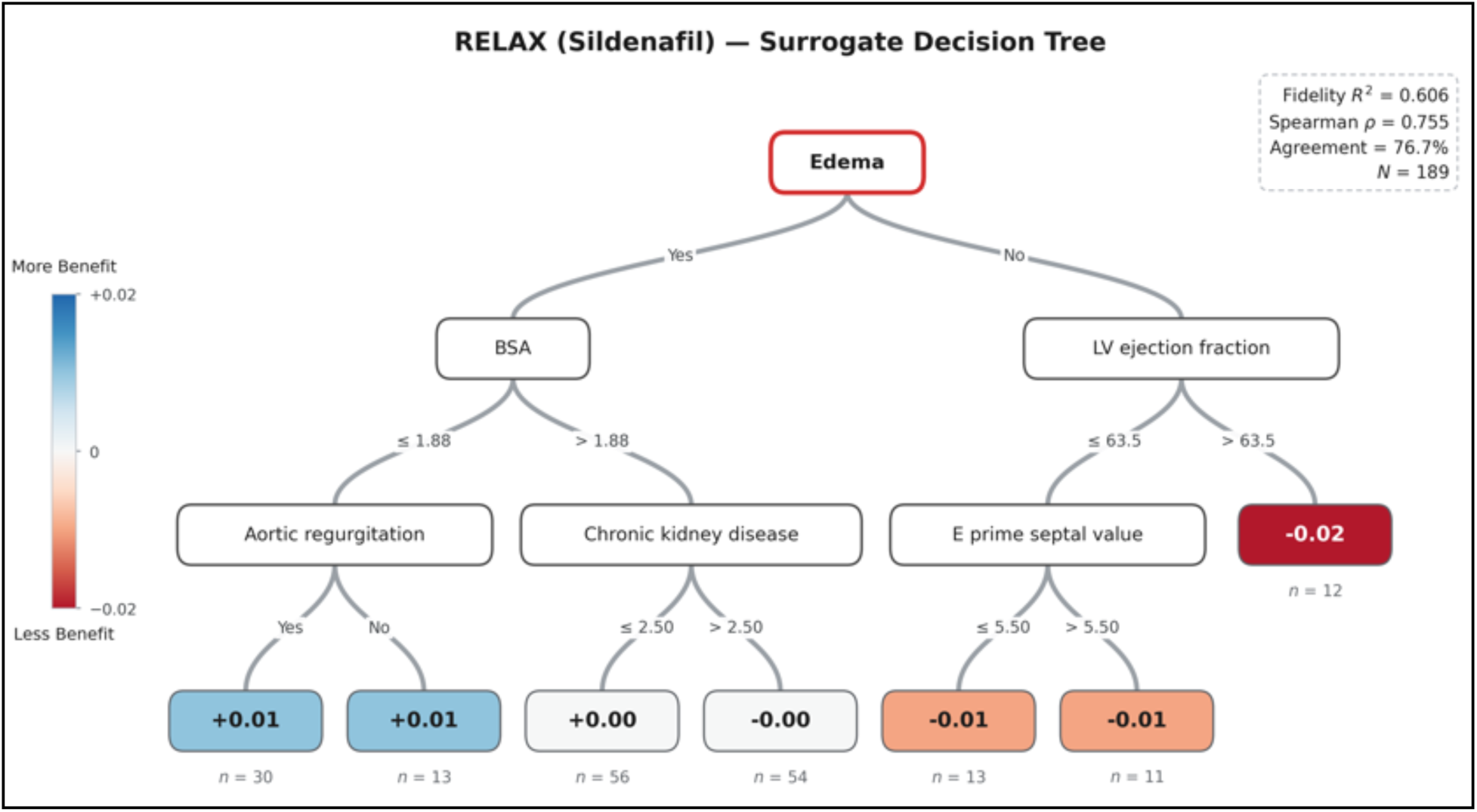
Surrogate decision tree approximating the RELAX-trained interaction model for sildenafil treatment effect prediction. Diabetes mellitus was the primary split. Among non-diabetic patients, the tree predicted harm across all branches: smaller body surface area (≤2.24 m²) was associated with progressively greater harm at higher BMI (≥30.6: ITE = −0.20), and larger BSA (>2.24 m²) was associated with the most negative ITE among patients without atrial fibrillation/flutter (−0.31). Among diabetic patients, the direction reversed: those with mitral regurgitation showed predicted benefit, particularly non-smokers (+0.30), whereas diabetic patients without mitral regurgitation showed modest harm (−0.17). Surrogate fidelity: R² = 0.540, Spearman ρ = 0.742, high/low agreement = 80.6%; N = 191.

## 4. Discussion

This study lays out a framework that separates prognostic from predictive phenotyping in HFpEF. The study illustrates that the conventional machine learning “responder” model (AUROC 0.76) does an excellent job in identifying patients who improves over time and demonstrates a prognostic value for hard clinical endpoints. Nonetheless, this approach captures only prognosis, and does not identify who specifically benefits from treatment. In contrast, evidence suggests that the proposed interaction-based ITE model isolates treatment-specific signals of the trials, revealing that the overall null results of the HFpEF trials mask meaningful and drug-specific treatment effect heterogeneity.

### The Prognostic–Predictive Distinction in HFpEF

Although the distinction between prognostic and predictive biomarkers is well established in fields such as oncology, the same cannot be said about HFpEF research. In this context, the results presented in this work show why that distinction is important.

The prognostic model predicted survival outcomes in both treatment and placebo arms. It did not generalize well to non-HFpEF heart failure populations. Importantly, it showed no significant treatment-by-subgroup interaction (p = 0.17). In other words, this model identifies participants who are sicker at baseline who may have more room to improve due to regression to the mean or natural fluctuation. Even though this can be clinically useful for risk stratification, it does not help decide *which drug to use* because it does not distinguish between pharmacological benefit and a favorable natural trajectory.

Nevertheless, this has implications for the broader HFpEF phenotyping literature. Many prior studies, especially those using clustering or supervised outcome prediction, have identified “responder phenotypes.” But few have formally tested whether those phenotypes show differential treatment effects, rather than just differential prognosis. Our findings suggest that many reported “responder” groups may reflect prognostic signal rather than true predictive biology. Without formal interaction testing, the difference cannot be identified.

### Drug-Specific Treatment Effect Modification

The interaction-based ITE model revealed drug-specific treatment-modifying phenotypes, supported by three convergent lines of evidence. First, each trial’s ITE model showed significant treatment-by-subgroup interaction only for its own drug (TOPCAT p=0.001; NEAT-HFpEF p=0.003; INDIE-HFpEF p=0.004). Second, ITE–outcome correlations were significant only within the trial’s own treatment arm. Third, the modifying variables were biologically coherent with each drug’s mechanism of action.

For spironolactone in TOPCAT, the dominant modifier was a cardiorenal phenotype with creatinine clearance <30, diabetes, BUN, oral hypoglycemic use, and bundle branch block as the most prominent variables. These variables are consistent with aldosterone antagonism. Spironolactone works through the renin–angiotensin–aldosterone system and has clear cardiorenal effects. Interestingly, creatinine clearance <30 was associated with greater benefit (positive interaction coefficient), that might reflect a subgroup in whom aldosterone-driven fibrosis plays a larger pathophysiologic role. Similarly, bundle branch block and QRS prolongation may reflect higher cardiac conduction system fibrosis, in whom aldosterone antagonism may exert beneficial effects.

For isosorbide mononitrate in NEAT-HFpEF, the key modifiers were pulse pressure, rales, and systolic blood pressure. Mechanistically, nitrates primarily reduce preload through venous vasodilation, consistent with the identified modifiers. Patients with high pulse pressure and rales may represent more advanced diastolic dysfunction or vascular stiffness. These patients also present high afterload (vascular stiffness) and volume, and without addressing it, preload reduction alone may be inadequate. In that setting, aggressive preload reduction might actually be counterproductive. This aligns with the trial’s overall finding that nitrate therapy reduced daily activity levels, suggesting that for many HFpEF patients, the hemodynamic trade-offs outweigh potential benefits.

For inorganic nitrite in INDIE-HFpEF, modifiers included a volume-overload and metabolic phenotype. Nitric oxide–mediated vasodilation may improve peripheral oxygen delivery and offer anti-inflammatory benefits, but in patients with severe congestion or metabolic dysfunction, that mechanism may be deletereous or insufficient to overcome underlying pathophysiology.

### Reinterpreting TOPCAT: Heterogeneity Hidden in a “Neutral” Trial

One of the most clinically striking findings comes from TOPCAT. The ITE model identified a +7.2 KCCQ-point differential treatment effect between predicted high- and low-ITE subgroups (p=0.001). In other words, within a trial considered overall neutral, one subgroup benefits meaningfully (TE = +2.4 KCCQ points) while another subgroup may actually be harmed (TE = −4.8 KCCQ points).

A 4.8 point decrement approaches the 5-point MCID threshold for KCCQ, which is considered clinically relevant. This could reframe the trial, and rather than “spironolactone has limited effect for HFpEF,” the narrative could be “spironolactone has differential effects that needs cautious consideration.” Patient-level phenotyping provides a more principled explanation for the heterogeneous results of this trial. The high-ITE group also presented lower rates of hospitalization and mortality (**Table 8**).

**Table 8.** Clinical Outcomes by Predicted Treatment Benefit Subgroup in TOPCAT (Treatment Arm). Patients receiving spironolactone were stratified by median split of the predicted individualized treatment effect (ITE) into Higher Predicted Benefit (n=855) and Lower Predicted Benefit (n=854) groups. The upper panel reports event counts and rates for each clinical endpoint with log-rank P values. The lower panel reports unadjusted hazard ratios (95% CI) from Cox proportional hazards regression, with the Higher Predicted Benefit group as reference. Across all five endpoints, patients classified as Lower Predicted Benefit experienced significantly higher event rates, with hazard ratios ranging from 1.40 for cardiovascular death to 2.29 for HF hospitalization

| Outcome, n (%) | Higher Predicted Benefit (n=855) | Lower Predicted Benefit (n=854) | Log-rank P |
| --- | --- | --- | --- |
| CV Death or HF Hospitalization | 117 (13.7%) | 197 (23.1%) | <0.001 |
| All-cause Death | 105 (12.3%) | 147 (17.2%) | 0.002 |
| Cardiovascular Death | 67 (7.8%) | 88 (10.3%) | 0.037 |
| All-cause Hospitalization | 313 (36.7%) | 446 (52.3%) | <0.001 |
| HF Hospitalization | 66 (7.7%) | 139 (16.3%) | <0.001 |
| **Unadjusted HR (95% CI)** |  |  | **Cox P** |
| CV Death or HF Hospitalization | 1.0 (Ref) | 1.85 (1.47-2.33)‡ | <0.001 |
| All-cause Death | 1.0 (Ref) | 1.49 (1.16-1.92)† | 0.002 |
| Cardiovascular Death | 1.0 (Ref) | 1.40 (1.02-1.92)* | 0.038 |
| All-cause Hospitalization | 1.0 (Ref) | 1.67 (1.45-1.93)‡ | <0.001 |
| HF Hospitalization | 1.0 (Ref) | 2.29 (1.71-3.07)‡ | <0.001 |

TOPCAT (Spironolactone) — Treatment arm only. Reference: Higher Predicted Benefit. \* $P<0.05$ ; † $P\leq 0.01$ ; ‡ $P\leq 0.001$ . Event counts: log-rank P. HR: Cox PH (Wald test).

### Relation to Prior Machine Learning Approaches in HFpEF

A study by Kresoja et al.^50^ used a sophisticated reiterative machine learning pipeline to identify spironolactone responders in the Aldo-DHF trial, which was later validated in TOPCAT. Their clustering design partially addresses the prognostic confounders by requiring differential improvement in E/e’ (variable used as their primary endpoint) between treatment and placebo within the responder cluster. Despite representing a major advancement towards novel approaches to analyzing the results of HFpEF trials, it is worth noting, however, that their model was trained to distinguish cluster membership rather than to model treatment-by-variable interactions directly, and the TOPCAT validation did not include a formal test of the treatment-by-subgroup interaction, making it is difficult to determine whether the reported difference between responders and non-responders is due to treatment effect modification.

Moreover, the variable profiles identified by their model are comprised predominantly of markers of general disease severity without a clear mechanistic link to aldosterone antagonism. In contrast, the treatment-modifying variables identified by our interaction-based model, such as creatinine clearance <30, diabetes mellitus, oral hypoglycemic use, and bundle branch block, are mapped onto the cardiorenal axis through which spironolactone exerts its effects^51,52^. This distinction is reinforced by our cross-trial specificity analysis, in which the TOPCAT model showed significant differential treatment effects only for spironolactone and failed to transfer to trials testing mechanistically distinct therapies. Fundamentally, while Kresoja’s approach identifies patients whose clinical profiles resemble those of treatment-arm improvers, our model directly isolates variables that modify the treatment effect relative to placebo.

Our work also differs from Thangaraj et al., whose model focuses on projecting TOPCAT-derived individualized treatment effects onto electronic health record (EHR) populations^53^. The question they attempted to answer is which real-world patients resemble the trial patients and could be potential responders. Although such an effort to achieve real-world translation underscores the need for ITE models that capture treatment-specific rather than prognostic signals, their model predicted that all EHR patients would benefit from spironolactone. This result suggests limited discriminative capacity for individualized treatment selection at this stage. Nevertheless, Thangaraj’s framework could serve as a valuable downstream application for our model. This framework has the potential to enable the projection of treatment-specific ITE estimates, such as those derived from our interaction-based model, onto real-world populations.

These findings carry direct implications for how HFpEF trials are designed and interpreted. Rather than relying solely on the average treatment effect to determine the therapeutic value, future trials should incorporate pre-specified analyses to determine whether a null overall result conceals meaningful benefit in identifiable subgroups. When pre-trial heterogeneity assessment using existing data suggests substantial treatment-effect variation, investigators should consider adaptive enrichment designs that stratify or restrict enrollment based on predicted benefit. The treatment-modifying variables identified in this study, particularly diabetes and renal function for spironolactone, are routinely collected at the point of care and could feasibly inform individualized prescribing if validated prospectively. More fundamentally, the consistent finding that drug-specific subgroups show both differential symptomatic response and differential long-term clinical outcomes suggests that meaningful progress toward HFpEF precision therapeutics may be possible using clinical variables already available in routine practice.

### Limitations

A few limitations need to be acknowledged. First, all analyses were retrospective. The treatment-modifying phenotypes we identified require prospective validation. Ideally, this should be performed in independent cohorts, or in enrichment-design clinical trials specifically powered to test interaction-based hypotheses.

Second, the smaller validation trials (RELAX, NEAT-HFpEF, INDIE-HFpEF; n = 191–215) have limited statistical power. This may explain the non-significant RELAX self-validation result (p = 0.317) and may also lead us to underestimate the true magnitude of treatment effect heterogeneity in these populations.

Third, differences in primary endpoints across trials make direct comparisons of effect sizes challenging. Although all endpoints used are clinically meaningful, they reflect different dimensions linked to functional status and symptom burden. These differences limit quantitative comparability across studies.

Finally, the relatively low R² values of the ITE models highlight an important reality: baseline clinical variables explain only part of the treatment effect heterogeneity in HFpEF. More granular data modalities (e.g., genomics, proteomics, metabolomics, or advanced imaging) may be necessary to achieve more precise and clinically actionable prediction of treatment effects.

## 5. Conclusions

Conventional responder analysis in HFpEF, which includes some machine learning approaches, is primarily designed to identify prognostic phenotypes in patients with favorable natural trajectories rather than patients who benefit from a specific drug. Interaction-based ITE models, in contrast, isolate treatment-specific, drug-mechanism–aligned signals and represent a promising approach to analyze treatment response in HFpEF trials. Leveraging the presented ITE model, we were able to detect signals related to cardiorenal variables that modify spironolactone response, hemodynamic variables that alter nitrate response, and volume-overload variables that change inorganic nitrite response. The cross-trial validation matrix, with diagonal significance only, supports mechanism specificity.

Clinically, the surrogate decision trees provide interpretable workflow for phenotype-guided treatment selection. In TOPCAT, for example, the ITE model distinguishes patients likely to benefit from spironolactone from those likely to experience net harm. This distinct group was obscured by the overall null result of the TOPCAT trial.

From a clinical trial-design perspective, the results of the proposed framework suggest that interaction-based enrichment strategies, such as enrolling patients predicted to have high ITE, may improve power and therapeutic gain in future HFpEF trials. In this context, separating prognosis from prediction is a step toward precision medicine in HFpEF.

## Data Availability

The study data access and analysis were completed entirely on the BioData Catalyst Platform under data use agreements through the HeartShare/AMP HF program and the National Institutes of Health's database of Genotypes and Phenotypes (dbGaP). Study data will not be shared but can be requested through these programs. Study datasets included clinical data from NHLBI-sponsored randomized trials (TOPCAT, RELAX, NEAT-HFpEF, and INDIE-HFpEF). Statistical and analytical code may be available on request.

## Acknowledgements

The HeartShare Program Next Generation Phenomics to Define HFpEF was supported through the following NHLBI grants: U54 HL160273 (Northwestern University Data Translation Center); U01 HL160279 (Northwestern University); U01 HL160277 (University of Pennsylvania); U01 HL160274 (University of California at Davis); U01 HL160226 (Mayo Clinic); U01 HL160272 (Wake Forest); U01 HL160278 (Massachusetts General Hospital) and through the FNIH Accelerating Medicines Partnership Heart Failure (AMP HF) Program [including RFP 2023-1345-001 (Johns Hopkins University)]. AHA 23SFRNCCS1052478, 23SFRNPCS1060482, and Research Award from the Rosenfeld Foundation (MC, DL); American College of Cardiology Foundation (MC). The authors also wish to acknowledge the contributions of the consortium working on the development of the NHLBI BioData Catalyst® (BDC) ecosystem.

## Disclaimer

The views expressed in this manuscript are those of the authors and do not necessarily represent the views of the National Heart, Lung, and Blood Institute; the National Institutes of Health; or the U.S. Department of Health and Human Services.

## References

1. Dunlay, S. M., Roger, V. L. & Redfield, M. M. Epidemiology of heart failure with preserved ejection fraction. Nat. Rev. Cardiol. 14, 591–602 (2017).

2. Savarese, G. et al. Global burden of heart failure: a comprehensive and updated review of epidemiology. Cardiovasc. Res. 118, 3272–3287 (2023).

3. Shahim, B., Kapelios, C. J., Savarese, G. & Lund, L. H. Global Public Health Burden of Heart Failure: An Updated Review. Card. Fail. Rev. 9, e11 (2023).

4. Borlaug, B. A. Evaluation and management of heart failure with preserved ejection fraction. Nat. Rev. Cardiol. 17, 559–573 (2020).

5. Shah, S. J. et al. Phenotype-Specific Treatment of Heart Failure With Preserved Ejection Fraction: A Multiorgan Roadmap. Circulation 134, 73–90 (2016).

6. Paulus, W. J. & Tschöpe, C. A novel paradigm for heart failure with preserved ejection fraction: Comorbidities drive myocardial dysfunction and remodeling through coronary microvascular endothelial inflammation. J. Am. Coll. Cardiol. 62, 263–271 (2013).

7. Santana, C., Cadeiras, M. & Liem, D. A. Multi-Organ Crosstalk in HFpEF: Reframing a Cardiac Syndrome as a widespread Systemic Disease. The Journal of Precision Medicine: Health and Disease 0, 100042 (2026).

8. Epelde, F. Heterogeneity in Heart Failure with Preserved Ejection Fraction: A Systematic Review of Phenotypic Classifications and Clinical Implications. Journal of Clinical Medicine 2025, Vol. 14, Page 4820 14, 4820 (2025).

9. Cuijpers, I. et al. Microvascular and lymphatic dysfunction in HFpEF and its associated comorbidities. Basic Research in Cardiology 2020 115:4 115, 39- (2020).

10. Deichl, A., Wachter, R. & Edelmann, F. Comorbidities in heart failure with preserved ejection fraction. Herz 2022 47:4 47, 301–307 (2022).

11. Anker, S. D. et al. Patient Phenotype Profiling in Heart Failure with Preserved Ejection Fraction to Guide Therapeutic Decision Making. A Scientific Statement of the Heart Failure Association, the European Heart Rhythm Association of the European Society of Cardiology, and …. Eur. J. Heart Fail. 25, 936–955 (2023).

12. Romero, E. et al. Clinical, Echocardiographic, and Longitudinal Characteristics Associated With Heart Failure With Improved Ejection Fraction. American Journal of Cardiology 211, 143–152 (2024).

13. Sharma, A. et al. Optimizing Foundational Therapies in Patients With HFrEF: How Do We Translate These Findings Into Clinical Care? JACC Basic Transl. Sci. 7, 504–517 (2022).

14. Wintrich, J. et al. Therapeutic approaches in heart failure with preserved ejection fraction: past, present, and future. Clinical Research in Cardiology 2020 109:9 109, 1079–1098 (2020).

15. Zakeri, R. & Cowie, M. R. Heart failure with preserved ejection fraction: controversies, challenges and future directions. Heart 104, 377–384 (2018).

16. Spertus, J. A. et al. Interpreting Population Mean Treatment Effects in the Kansas City Cardiomyopathy Questionnaire: A Patient-Level Meta-Analysis. JAMA Cardiol. 10, 32 (2025).

17. Royston, P., Altman, D. G. & Sauerbrei, W. Dichotomizing continuous predictors in multiple regression: a bad idea. Stat. Med. 25, 127–141 (2006).

18. Southworth, M. R., Psotka, M. A., Pomeroy, J. E. & Stockbridge, N. L. Responder Analyses Can Be Misleading. J. Am. Coll. Cardiol. 85, 196–198 (2025).

19. Shalit, U., Johansson, F. D. & Sontag, D. Estimating individual treatment effect: generalization bounds and algorithms. 3076–3085 Preprint at https://proceedings.mlr.press/v70/shalit17a.html (2017).

20. Mcmurray, J. J. V & O’connor, C. Lessons from the TOPCAT Trial. n engl j med 370 doi:10.1056/NEJMe1401231.

21. Wang, H. et al. Sildenafil Treatment in Heart Failure With Preserved Ejection Fraction: Targeted Metabolomic Profiling in the RELAX Trial. JAMA Cardiol. 2, 896–901 (2017).

22. Honigberg, M. C. et al. Sex Differences in Exercise Capacity and Quality of Life in Heart Failure With Preserved Ejection Fraction: A Secondary Analysis of the RELAX and NEAT-HFpEF Trials. J. Card. Fail. 26, 276–280 (2020).

23. Borlaug, B. A. et al. Effect of Inorganic Nitrite vs Placebo on Exercise Capacity Among Patients With Heart Failure With Preserved Ejection Fraction: The INDIE-HFpEF Randomized Clinical Trial. JAMA 320, 1764–1773 (2018).

24. HeartShare Consortium Submitter. HeartShare - Extant Datasets - Harmonized Clinical Trials Collection. https://www.ncbi.nlm.nih.gov/projects/gap/cgi-bin/collection.cgi?study_id=phs003989.v1.p1 (2025).

25. Savill, P. Spironolactone in heart failure with preserved ejection fraction. Practitioner 258, 10 (2014).

26. Lindman, B. R. et al. Cardiovascular Phenotype in Patients with Heart Failure and Preserved Ejection Fraction with or without Diabetes: A RELAX Trial Ancillary Study. J. Am. Coll. Cardiol. 64, 541 (2014).

27. Zakeri, R. et al. Nitrate’s effect on activity tolerance in heart failure with preserved ejection fraction trial rationale and design. Circ. Heart Fail. 8, 221–228 (2015).

28. Reddy, Y. N. V. et al. Inorganic Nitrite Delivery to Improve Exercise Capacity in Heart Failure with Preserved Ejection Fraction (INDIE-HFpEF): Rationale and Design. Circ. Heart Fail. 10, e003862 (2017).

29. Caraballo, C. et al. Clinical implications of the New York heart association classification. ahajournals.org 8, (2019).

30. Mathai, S. C., Puhan, M. A., Lam, D. & Wise, R. A. The Minimal Important Difference in the 6-Minute Walk Test for Patients with Pulmonary Arterial Hypertension. 10.1164/rccm.201203-0480OC 186, 428–433 (2012).

31. National Heart, L. and B. I. N. I. of H. U. S. D. of H. and H. S. The NHLBI BioData Catalyst. 10.5281/ZENODO.3822858 doi:10.5281/ZENODO.3822858.

32. Wall, M. E., Rechtsteiner, A. & Rocha, L. M. Singular Value Decomposition and Principal Component Analysis. A Practical Approach to Microarray Data Analysis 91–109 (2003) doi:10.1007/0-306-47815-3_5.

33. Ponce-Bobadilla, A. V., Schmitt, V., Maier, C. S., Mensing, S. & Stodtmann, S. Practical guide to SHAP analysis: Explaining supervised machine learning model predictions in drug development. Clin. Transl. Sci. 17, (2024).

34. Cohen, J. B. et al. Clinical Phenogroups in Heart Failure With Preserved Ejection Fraction: Detailed Phenotypes, Prognosis, and Response to Spironolactone. JACC Heart Fail. 8, 172–184 (2020).

35. Swaminathan, K. et al. Spironolactone for poorly controlled hypertension in type 2 diabetes: conflicting effects on blood pressure, endothelial function, glycaemic control and hormonal profiles. Diabetologia 51, 762–768 (2008).

36. Davies, J. I., Band, M., Morris, A. & Struthers, A. D. Spironolactone impairs endothelial function and heart rate variability in patients with type 2 diabetes. Diabetologia 47, 1687–1694 (2004).

37. de Denus, S. et al. Spironolactone Metabolites in TOPCAT - New Insights into Regional Variation. N. Engl. J. Med. 376, 1690–1692 (2017).

38. Pfeffer, M. A. et al. Regional variation in patients and outcomes in the treatment of preserved cardiac function heart failure with an aldosterone antagonist (TOPCAT) trial. Circulation 131, 34–42 (2015).

39. Bristow, M. R. et al. Detection and Management of Geographic Disparities in the TOPCAT Trial: Lessons Learned and Derivative Recommendations. JACC Basic Transl. Sci. 1, 180–189 (2016).

40. Pfeffer, M. A. & Claggett, B. Behind the Scenes of TOPCAT — Bending to Inform. NEJM Evidence 1, (2022).

41. Patel, J. N. & Shah, S. J. Inorganic vs. organic nitrates for heart failure with preserved ejection fraction: it’s not all in your head! Eur. J. Heart Fail. 19, 1516–1519 (2017).

42. Cai, Z. et al. The NO-cGMP-PKG Axis in HFpEF: From Pathological Mechanisms to Potential Therapies. Aging Dis. 14, 46 (2023).

43. Borlaug, B. A. et al. Effect of Inorganic Nitrite vs Placebo on Exercise Capacity Among Patients With Heart Failure With Preserved Ejection Fraction: The INDIE-HFpEF Randomized Clinical Trial. JAMA 320, 1764–1773 (2018).

44. Andersson, K. E. PDE5 inhibitors – pharmacology and clinical applications 20 years after sildenafil discovery. Br. J. Pharmacol. 175, 2554–2565 (2018).

45. Balarini, C. M. et al. Sildenafil restores endothelial function in the apolipoprotein E knockout mouse. J. Transl. Med. 11, 3 (2013).

46. Lundberg, J. O., Weitzberg, E. & Gladwin, M. T. The nitrate-nitrite-nitric oxide pathway in physiology and therapeutics. Nat. Rev. Drug Discov. 7, 156–167 (2008).

47. Münzel, T., Daiber, A. & Gori, T. Nitrate therapy: New aspects concerning molecular action and tolerance. Circulation 123, 2132–2144 (2011).

48. Thomas, G. R., DiFabio, J. M., Gori, T. & Parker, J. D. Once Daily Therapy With Isosorbide-5-Mononitrate Causes Endothelial Dysfunction in Humans: Evidence of a Free-Radical–Mediated Mechanism. J. Am. Coll. Cardiol. 49, 1289–1295 (2007).

49. Oelze, M. et al. Chronic therapy with isosorbide-5-mononitrate causes endothelial dysfunction, oxidative stress, and a marked increase in vascular endothelin-1 expression. Eur. Heart J. 34, 3206– 3216 (2013).

50. Kresoja, K.-P. et al. Treatment response to spironolactone in patients with heart failure with preserved ejection fraction: a machine learning-based analysis of two randomized controlled trials. EBioMedicine 96, 104795 (2023).

51. Zannad, F. et al. Eplerenone in patients with systolic heart failure and mild symptoms. N. Engl. J. Med. 364, 11–21 (2011).

52. Bakris, G. L. et al. Effect of Finerenone on Chronic Kidney Disease Outcomes in Type 2 Diabetes. New England Journal of Medicine 383, 2219–2229 (2020).

53. Thangaraj, P. M. et al. Computational Phenomapping of Randomized Clinical Trials to Enable Assessment of their Real-world Representativeness and Personalized Inference. 10.1101/2024.05.15.24306285 doi:10.1101/2024.05.15.24306285.

